# Identifying individuals with rare disease variants by inferring shared ancestral haplotypes from SNP array data

**DOI:** 10.1101/2023.12.20.23300328

**Authors:** Erandee Robertson, Bronwyn E Grinton, Karen L Oliver, Liam G Fearnley, Michael S Hildebrand, Lynette G Sadleir, Ingrid E Scheffer, Samuel F Berkovic, Mark F Bennett, Melanie Bahlo

## Abstract

We describe FoundHaplo, a novel identity-by-descent algorithm designed to identify individuals with known, untyped, disease-causing variants using only SNP array data. FoundHaplo leverages knowledge of shared disease haplotypes for inherited disease-causing variants to identify individuals who share the disease haplotype and are, therefore, likely to carry the rare (MAF<0.01) variant. We performed a simulation study to evaluate the performance of FoundHaplo across 33 known disease-harbouring loci. We demonstrated the ability of FoundHaplo to infer the presence of two rare (MAF<0.01) pathogenic variants, *SCN1B* c.363C>G (p.Cys121Trp) and *WWOX* c.49G>A (p.E17K), which can cause mild dominant and severe recessive epilepsy respectively, in two large cohorts including 1,573 individuals with epilepsy from the Epi25 cohort and 468,481 individuals from the UK Biobank. We demonstrate that FoundHaplo performs substantially better at inferring the presence of these variants than existing genome-wide imputation approaches. FoundHaplo is a valuable, low-cost screening tool that can be applied to search SNP genotyping array data for disease-causing variants with known founder effects based on shared disease haplotypes. FoundHaplo is available at https://github.com/bahlolab/FoundHaplo.

## INTRODUCTION

Detecting disease-causing variants (DCVs) is essential for identifying individuals at high-risk for disease ^1,2^, enabling appropriate patient care. Many pathogenic variants observed in unrelated individuals or families may result from the variant arising independently or may be the result of a variant being inherited from a common ancestor. This leads to shared core haplotypes among carriers, a phenomenon known as a founder effect ^3–10^. DCVs with founder events thus have an associated disease haplotype inherited from the common ancestor, shared by all the descendant variant carriers in subsequent generations ^3–10^. Pathogenic variants initially thought to be recurrent have been able to be reclassified as inherited based on haplotype sharing from a common ancestral founder ^6,10^. Shared haplotypes inherited from a common ancestor are defined as being identical by descent (IBD). Haplotypes shared by carriers of the DCV decrease in size over generations due to recombination events ^3–10^. Regardless of the time elapsed since the original founder event, an IBD segment persists in current-day descendants carrying the DCV. This suggests that detecting associated disease haplotypes through an IBD approach can also infer the presence of these inherited DCVs.

Founder events for many DCVs have been previously described. Table S1 lists an illustrative set of inherited genetic disorders with reported founder effects. Some genetic disorders show evidence of multiple founder events, each with its own unique haplotype. Examples of founder events include the Huntington’s disease repeat expansion, which displays multiple founder events ^7,11–13^, the *CFTR* p.F508del Cystic fibrosis-causing variant ^14^ and the *GOSR2* p.G144W progressive myoclonus epilepsy-causing variant ^15^. In general, the population frequency of these highly or fully penetrant DCVs is low, with a minor allele frequency (MAF) <0.01 or 1%, typically leading to rare diseases. This low MAF typically leads to these variants being excluded in genome-wide association studies.

Most published IBD methods seek to identify genome-wide IBD tracts rather than directly screening individuals for DCVs ^16–22^. They do not make use of DCV haplotype information. DRIVE is a recent IBD tool developed to cluster carriers of DCVs to try and identify additional DCV carriers using existing genome-wide IBD methods ^23^.

Imputation platforms such as The Michigan Imputation Server (MIS) ^24^ and the TOPMed server ^25^ utilise linkage disequilibrium (LD) for genome-wide imputation of millions of variants not directly genotyped by SNP arrays. However, these rely on having DCV haplotypes in their reference databases, which is often not the case. With lower MAF, these imputation servers also perform more poorly. Hence, these approaches are not designed to identify rare DCVs (MAF<0.01).

Here, we introduce FoundHaplo, a novel IBD-based tool, developed using a first-order Hidden Markov Model (HMM), to detect DCVs with known founder effects from shared disease haplotypes requiring only SNP genotyping array data. FoundHaplo leverages its inference on pre-existing information of the location and haplotype of the DCV. This tool is particularly relevant given the widespread use of SNP genotyping arrays in genome-wide association studies (GWAS) to patient cohorts and large biobanks such as the UK Biobank (UKBB) ^26^, where many individuals are genotyped but not sequenced due to the relatively high costs of genome sequencing in contrast to SNP genotyping arrays or due to lack of remaining biospecimens suitable for sequencing ^27,28^. Even though SNP genotyping arrays are cost-effective and commonly available, many DCVs are not captured directly on SNP arrays. FoundHaplo addresses the gap in identifying DCVs not directly SNP genotyped or imputable with existing tools due to their low MAF and lack of representation in large databases leveraged for imputation^29–33^.

We perform a comprehensive simulation study to demonstrate the performance of FoundHaplo under single and multiple founder effects and then apply the algorithm to identify DCVs in cohorts, including the UKBB ^26^, demonstrating that FoundHaplo is a useful, low-cost screening tool which could be applied to bespoke catalogues of DCV haplotypes to identify individuals that merit further sequencing.

## MATERIAL AND METHODS

### FoundHaplo HMM

The FoundHaplo HMM aims to differentiate between random haplotype sharing and IBD between a known disease haplotype and a test individual in the vicinity of a DCV in a hypothesis-testing framework to infer the presence of a DCV in an individual’s phased and imputed genotyping data. The null hypothesis (H0) asserts no IBD between the individual’s haplotypes and the disease haplotype, indicating no common founder inheritance of the DCV. The alternative hypothesis (H1) suggests at least one haplotype presents evidence of IBD with the disease haplotype, indicating inheritance from a common founder. The FoundHaplo HMM, focusing on biallelic SNPs, models IBD to determine the hidden IBD state, discerning between no IBD (0) and IBD (1) based on the observed reference or alternate (0 or 1) alleles. HMMs in FoundHaplo replace the typical “waiting time” by the genetic map distance in Morgans from a known DCV locus to the next recombination event. With unknown IBD sharing boundaries, the algorithm starts Markov chains at the DCV locus and extends in opposite directions, comprising two Markov chains as illustrated in Figure 1.

**Figure 1.**
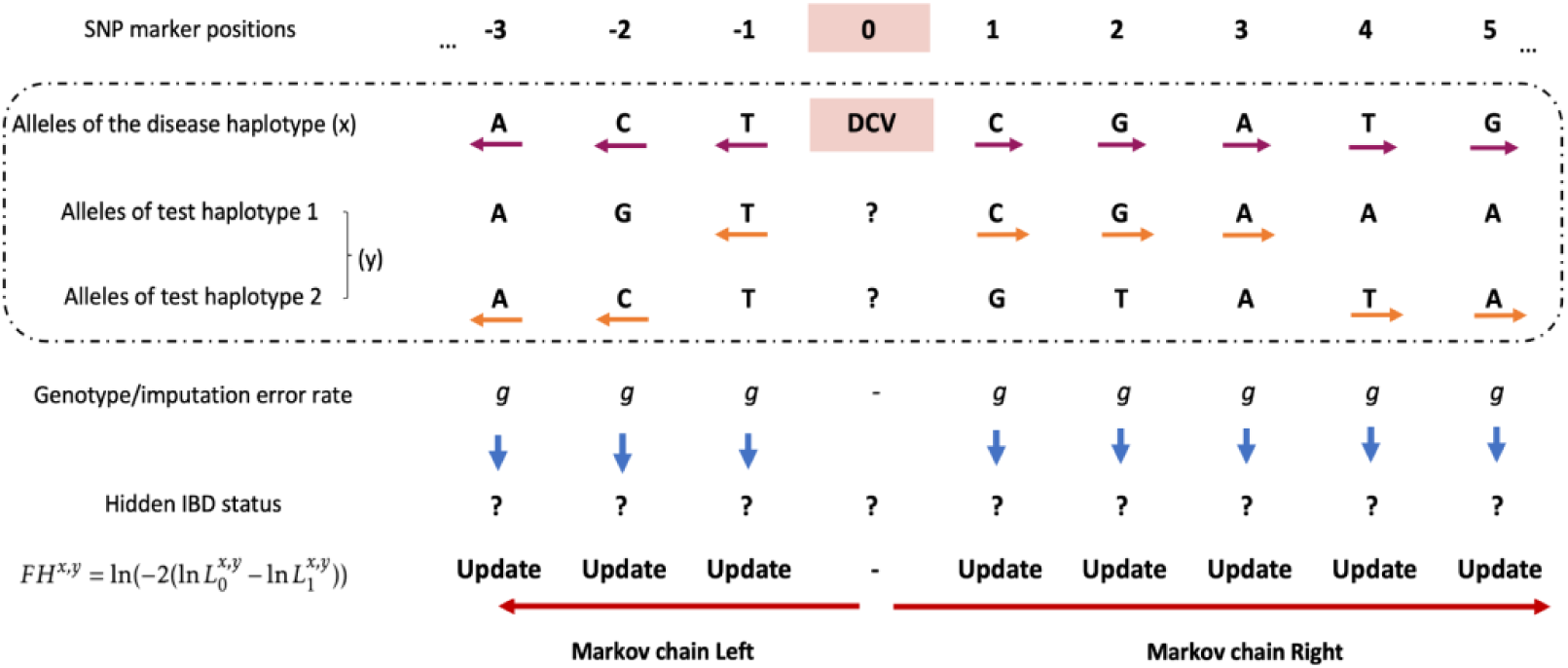
Calculating the *FH* score for a disease-test pair based on the HMM approach. IBD status is unknown at all the SNP markers, making IBD segment boundaries unknown. We test a disease haplotype against an individual’s two haplotypes, observing alleles surrounding the DCV, and moving from the DCV to the left and right.

The FoundHaplo algorithm calculates the log-likelihood ratio (*LLR*) of IBD versus non-IBD at each genetic marker surrounding the DCV (denoted by marker 0) for a disease haplotype and a test pair of imputed haplotypes. The likelihood of IBD is encapsulated in the FoundHaplo (*FH)* score, defined as ln(*LLR*) (Supplemental material and methods).

The algorithm updates the likelihood of hidden IBD (L0 for the null and L1 for the alternate hypothesis) in the *FH* score. It accounts for a fixed rate of genotype and imputation errors, and switches between the test individual’s two haplotypes to handle phasing errors.

Genotype and imputation errors are indistinguishable in this model and are treated similarly, assumed to occur at a fixed, uniform rate (*g*) of 1% across the genome ^34,35^ (Figure 1, Figure S1 and Supplemental material and methods). Genotype markers with missing values are excluded from the analysis.

To propagate the *LLR*, the FoundHaplo algorithm switches to the alternate haplotype of the test individual when haplotype sharing ceases on the current one, as depicted by orange arrows in Figure 1. This approach captures potential sharing on the other haplotype and accommodates block phasing errors introduced by LD-based phasing tools. The Markov chains end when sharing around the DCV between the disease haplotype and test individual stops (Supplemental material and methods). When multiple known disease haplotypes for a single disease exist, the *FH* score is determined as the maximum of individual *FH* scores across all available disease haplotypes for that variant (Supplemental material and methods).

### Using empirical p-values for *FH* score evaluation

In the FoundHaplo algorithm, while the log-likelihood ratio test statistics under the null hypothesis are theoretically asymptotically chi-square distributed ^36^, the actual distribution deviates due to linkage disequilibrium (Figure S5). Therefore, the significance of the FoundHaplo statistic is assessed using the empirical distribution of *FH* scores from a control population. A test individual is identified as having IBD sharing with a disease haplotype if their *FH* score exceeds a critical threshold, set based on the 99^th^ percentile of the *FH* score distribution in a control cohort of the same ancestral population as the test cohort, typically using data from the 1000 Genomes phase 3 haplotypes ^37^.

### Inputs required by FoundHaplo

FoundHaplo algorithm relies on two parameters: minor allele frequencies (MAF) of genetic markers and recombination frequency between markers. Ancestry-agnostic recombination rates for the human genome are commonly used in IBD algorithms ^5,9,22^. Recombination rates in FoundHaplo are calculated from the genetic maps available from the HapMap project ^38,39^. We recommend using the MAF of the relevant ancestry of the test cohort taken from the gnomAD database ^40,41^ since MAF varies by ancestral population ^24,32,33,37^. The algorithm does not incorporate LD since IBD segments are typically larger than the length of LD blocks.

FoundHaplo requires phased genotype data for both test individuals and disease haplotypes. To enhance the limited variant coverage of SNP array data, imputation is used to increase marker density. Imputed markers with R-squared ⩾0.3 are retained for FoundHaplo analysis in line with standard quality measures ^24,42,43^. Both imputation and phasing can be performed together using LD-based genome-wide imputation and phasing tools or servers such as the Michigan Imputation Server (MIS) ^24^ or TOPMed server ^25^.

FoundHaplo requires accurately phased disease haplotypes, best achieved through pedigree-phasing with another confirmed carrier of the same DCV from the same family. This avoids errors from LD-based genome-wide phasing, which only resolves phasing to LD block resolution. Additionally, individuals with long homozygosity regions due to related parents can be used as the source of recessive disease haplotypes, as the homozygosity tracts are often much longer than any shared IBD tracts.

The FoundHaplo algorithm, available as an R package with a disease haplotype database schema, is freely available from https://github.com/bahlolab/FoundHaplo.

Researchers can create and manage their own database instances of disease haplotypes, maintaining data confidentiality while running FoundHaplo. A detailed mathematical derivation of the algorithm is provided in the Supplemental material and methods.

### Simulation study

We performed a simulation study for 33 DCVs to evaluate the performance of the FoundHaplo algorithm using 503 unrelated individuals with European ancestry from the 1000 Genomes Project phase 3 dataset ^37^. Most of the DCVs we simulated are located at known repeat expansion loci (Table S3). Repeat expansions are rare, often inherited with known founder effects ^3,4,8,44^ and cannot be detected using SNP genotyping array data. They are, therefore, an excellent candidate set of diseases to demonstrate the utility of FoundHaplo. The simulation study investigated: (i) single founder effects, where multiple different versions of a single disease haplotype are found in the present time that are all distantly related and descended from a single ancestor, and (ii) multiple founder effects, where the same DCV has arisen independently in multiple unrelated founders, resulting in multiple unique haplotypes (Figure S2). Test cohorts were constructed by designating a fraction of individuals as cases. Each case was simulated by replacing the haplotype spanning the DCV locus with a randomly selected disease haplotype to simulate the presence of an inherited DCV. Cases were simulated to share different sizes of the disease haplotypes (0.5, 1, 2 and 5 cM) surrounding the DCV to simulate pairs of individuals with varying times to the most recent common ancestor (MRCA) (Figure S2). The rest of the test cohort remained unchanged, acting as controls. In our simulations, we introduced genotype and imputation errors by altering 1% of marker alleles genome-wide. Additionally, we simulated phasing errors in all individuals (except those used to derive the disease haplotypes) by switching blocks of adjacent marker alleles to the alternate haplotype with a rate of one switch per 20.05 Mbp ^45^.

For each of the 33 simulated disease loci, we created ten founder scenarios, generating ten disease haplotypes and 50 cases (5 per disease haplotype) for each scenario. This resulted in 1,320 simulation datasets encompassing both single and multiple founder effects with varying sharing lengths (Table S3 and Algorithm S2).

### Detecting the *SCN1B* c.363C>G and *WWOX* c.49G>A rare epilepsy variants

FoundHaplo was used to predict carriers of the *SCN1B* c.363C>G (p.Cys121Trp) and *WWOX* c.49G>A (p.E17K) rare variants in two cohorts. *SCN1B* c.363C>G has a MAF of 0.01047% in gnomAD ^40,41^ and causes autosomal dominant genetic epilepsy with febrile seizures plus (OMIM: 604233) ^46–49^. *WWOX* c.49G>A has a MAF of 0.01037% in gnomAD ^40,41^ and causes autosomal recessive developmental and epileptic encephalopathy (OMIM: 616211) ^50–52^.

Cohort 1 consisted of 1,573 individuals with different types of epilepsy recruited in Australia or New Zealand as part of the international Epi25 study ^53^. Cohort 2 is the UKBB cohort (n=468,481) accessed through project ID 36610 ^26^. Both cohorts, primarily of European ancestry and with WES and SNP genotyping data (Supplemental Material and Methods), identified two individuals in the Epi25 cohort and 171 individuals in the UKBB cohort who carried the *SCN1B* c.363C>G variant and 172 individuals in the UKBB who carried the *WWOX* c.49G>A variant.

For FoundHaplo analysis, five *SCN1B* c.363C>G and three *WWOX* c.49G>A disease haplotypes were created using duo and trio genotype data of eight different families (Supplemental Material and Methods). The European (EUR) cohort of 1000 Genomes Phase 3 ^37^ was used as the control cohort when using FoundHaplo. None of the samples in the EUR cohort of the 1000 Genomes data carried either of the two variants.

FoundHaplo predictions were computed using critical values at the 99, 99.5 and 99.8 percentiles from the 1000 Genomes data. The algorithm’s effectiveness was evaluated using the area under the precision-recall (PR) curve (AUPRC), appropriate for imbalanced datasets ^54,55^. The performance of a random classifier of a PR curve can be evaluated with the baseline rate, which is the ratio of positives to the total cohort size^54,55^.

### Performance comparison to DRIVE

We compared FoundHaplo with DRIVE ^23^, the only other known algorithm capable of detecting individuals with DCVs using haplotypes. DRIVE’s performance was evaluated for identifying carriers of *SCN1B* c.363C>G and the *WWOX* c.49G>A variants in the Epi25 cohort. To run the DRIVE algorithm, known carriers (five *SCN1B* c.363C>G and three *WWOX* c.49G>A carriers) of these variants were merged into the Epi25 dataset in two separate analyses for the two DCVs. DRIVE assessed if the individuals in the Epi25 cohort carrying these DCVs clustered with the original known carriers (Supplemental material and methods). We did not run DRIVE on the UKBB cohort (n=468,481) as the run time required for such a large number of samples was not feasible.

Briefly, DRIVE uses the output from existing genome-wide IBD detection tools and identifies clusters of test individuals containing individuals already known to harbour the DCV who pairwise share an IBD segment overlapping a locus of interest ^23^.

This study was approved by the Austin Health Human Research Ethics Committee. Informed consent was obtained and archived from all participants or their legal guardian. Research was approved by the Human Research Ethics Committee at The Walter and Eliza Hall Institute of Medical Research (G20/01, 17/09LR).

## RESULTS

### Simulation study

We performed a simulation study using the European cohort of the 1000 Genomes phase 3 data to test FoundHaplo’s ability to identify shared disease haplotypes. The performance in distinguishing simulated cases from controls was assessed under a variety of settings, including varying shared haplotype sizes, number of disease haplotypes, and DCV loci. The performance was evaluated for critical value percentiles of 99, 99.5 and 99.8.

Figure 2 displays FoundHaplo’s sensitivity at the default 99^th^ percentile, measured by its ability to identify simulated cases sharing an ancestor with the disease haplotypes in use. The algorithm showed 81% sensitivity for single founder effects and 69% for multiple founder effects with a shared length of at least 2 cM, maintaining an empirical false positive rate of 1%. Sensitivity is higher for single founder effects due to having a shared core ancestral haplotype. The sensitivity increases with larger simulated shared segments that can be better differentiated from the general population, indicative of a more recent common ancestor. Variations in performance across disease loci may arise from the frequency of chosen disease haplotypes in simulations or differing LD levels at certain loci.

**Figure 2.**
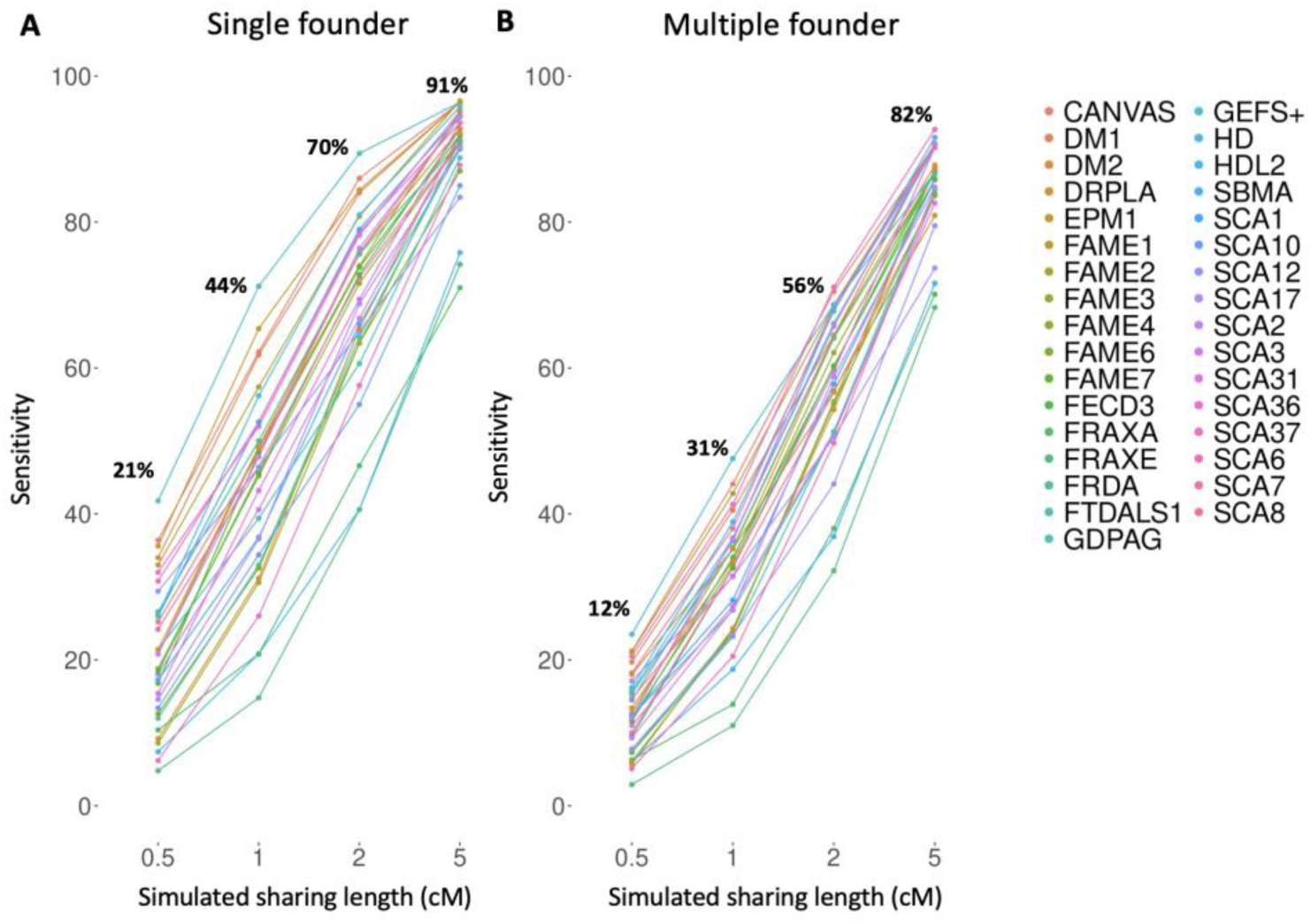
Sensitivity of the FoundHaplo algorithm for 33 simulated disease loci at the default 99^th^ percentile critical value. The sensitivity for (A) single founder effects and (B) multiple founder effects were calculated based on the ability to correctly predict simulated cases that have a common ancestor with the disease haplotypes in use.

Figure 3 demonstrates the variation in sensitivity and area under the PR curve (AUPRC) with increasing numbers of disease haplotypes. Sensitivity and AUPRC were calculated based on accurately predicting all simulated cases in each test cohort. The AUPRC of a random classifier in each simulation is 0.1 (50 cases/492 total cohort size)(black horizontal lines in Figure 3 C-D (Algorithm S2)).

**Figure 3.**
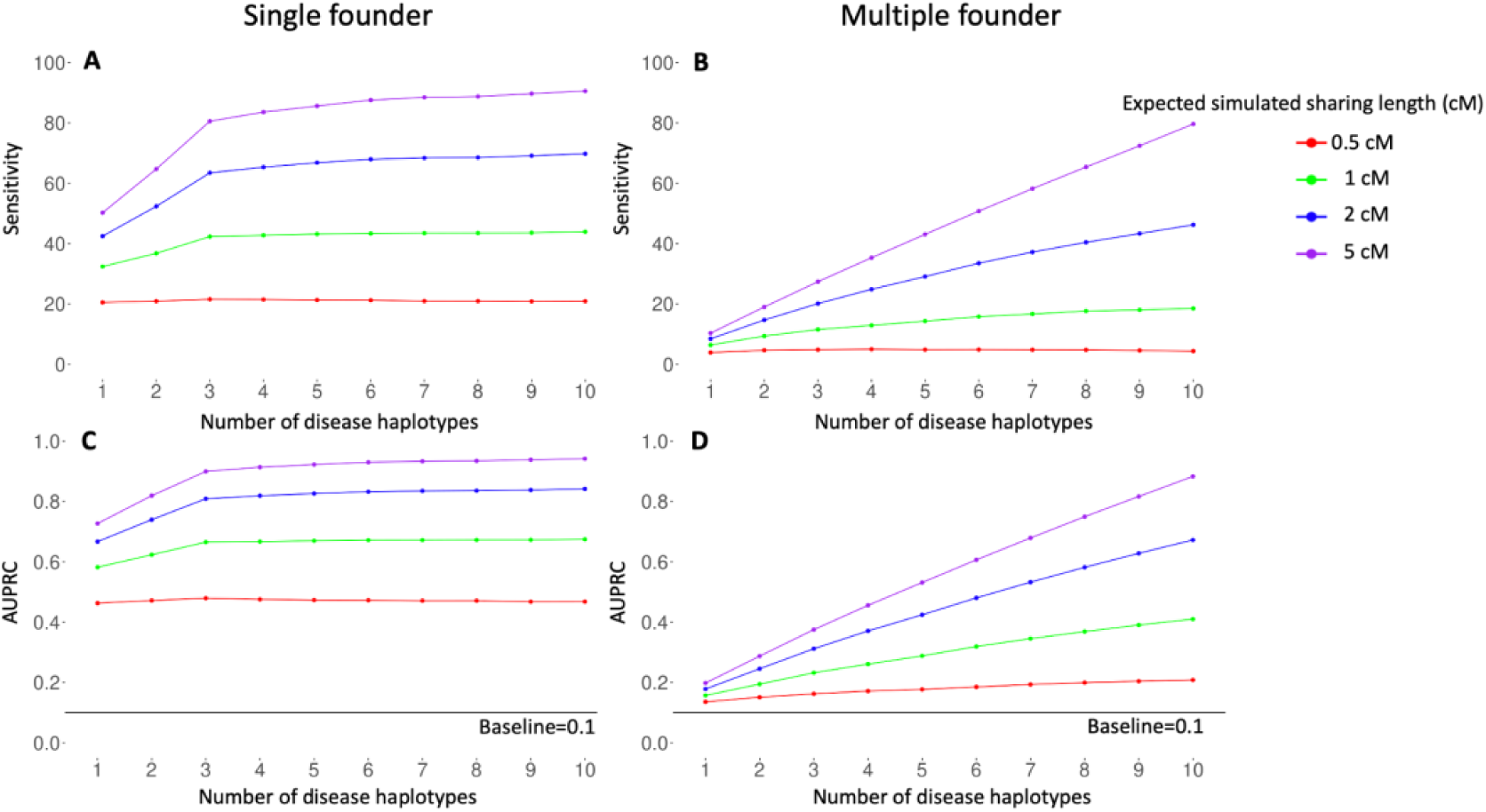
Performance of the FoundHaplo algorithm based on sensitivity and AUPRC by the number of disease haplotypes used averaged for all 33 simulated disease loci. The (A) sensitivity for single founder effects, (B) sensitivity for multiple founder effects, (C) AUPRC for single founder effects and (D) AUPRC for multiple founder effects were calculated based on the ability to correctly predict all the simulated cases in simulated test cohorts. The AUPRC of a random classifier in each simulation is 0.1 and is shown in black horizontal lines in (C) and (D).

Sensitivity and AUPRC are greater for single founder effects in FoundHaplo. The algorithm can not identify shared haplotypes without a common ancestor with known disease haplotypes, leading to lower sensitivity in multiple founder effects with fewer disease haplotypes. This illustrates how having more unique disease haplotypes enhances the performance, especially for DCVs with multiple founder effects, as shown in Figures 3B and D.

### Detecting individuals with the *SCN1B* c.363C>G and *WWOX* c.49G>A rare epilepsy variants

The original five *SCN1B* c.363C>G (p.Cys121Trp) carriers shared a core haplotype of 4.1 cM around the *SCN1B* c.363C>G variant, and the three *WWOX* c.49G>A (p.E17K) carriers shared a core haplotype of 3.9 cM around the *WWOX* c.49G>A variant, suggesting a common ancestor between the families for each of the two variants (Figures S10 and S11). A common ancestor for the *SCN1B* c.363C>G variant has already been identified by Grinton et al ^6^, and here we demonstrate evidence of a founder effect for the *WWOX* c.49G>A (p.E17K) variant.

Analysis of WES data identified two individuals in the Epi25 cohort and 171 in the UKBB cohort carrying the *SCN1B* c.363C>G variant and 172 individuals in the UKBB cohort carrying the recurrent *WWOX* c.49G>A variant. None of the Epi25 individuals were identified to carry the *WWOX* c.49G>A variant. We compared the disease haplotypes for these variants with the confirmed carriers based on WES analysis in the UKBB and Epi25 cohorts. All of the 178 *SCN1B* c.363C>G carriers shared a core haplotype of 55 kbp. Among carriers in the Epi25 and the UKBB, a minimum pairwise sharing of 63 kbp, a median of 4,650 kbp and a maximum of 12,321 kbp was observed. All of the 175 *WWOX* c.49G>A carriers shared a core haplotype of 157 kbp. Among carriers in the UKBB, a minimum pairwise sharing of 540 kbp, a median of 1,762 kbp and a maximum of 10,000 kbp was observed (Figures S10 and S11). This suggests that all the carriers have a common ancestor for each of the two variants. The two Epi25 carriers shared the shortest genomic region with the core haplotype (63 kbp and 373 kbp) around the *SCN1B* c.363C>G locus, implying that they are more distantly related compared to the rest of the carriers.

The distribution of the *FH* scores in the Epi25 (n=1,573) and the UKBB cohorts (n=468,481) and in their respective control cohorts for the constructed *SCN1B* c.363C>G and *WWOX* c.49G>A disease haplotypes are shown in Figure 4. Non-carriers above the 99^th^ percentile critical value are shown in black for the Epi25 but not for the UKBB cohort, due to the large number of individuals (n=4,685) at this percentile under the null hypothesis alone. Using the 99^th^ percentile, FoundHaplo predicted both *SCN1B* c.363C>G variant carriers and 13 non-carriers (100% sensitivity and 0.8% false positive rate) in the Epi25 cohort. There are no *WWOX* c.49G>A carriers in the Epi25 cohort, but FoundHaplo identified 23 non-carriers (1.5% false positive rate). FoundHaplo predicted 166 *SCN1B* c.363C>G variant carriers (97% sensitivity and 2% false positive rate) and 167 *WWOX* c.49G>A carriers correctly (97% sensitivity and 0.9% false positive rate) in the UKBB cohort.

**Figure 4.**
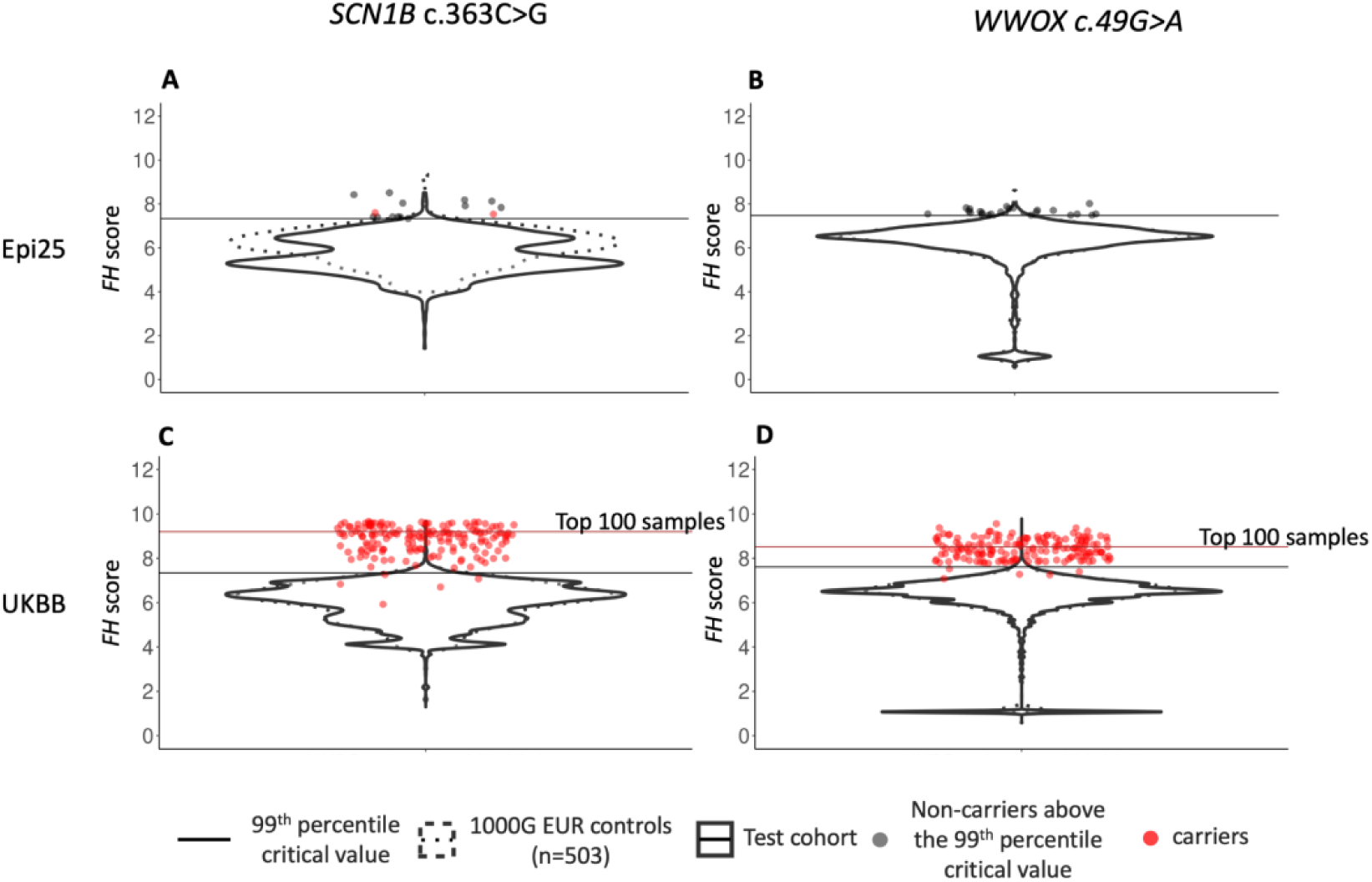
Distribution of *FH* scores in the Epi25 cohort and the UKBB cohort. Two disease variants are shown: (A) *SCN1B* c.363C>G variant in the Epi25 cohort, (B) *WWOX* c.49G>A variant in the Epi25 cohort, (C) *SCN1B* c.363C>G variant in the UKBB cohort and (D) *WWOX* c.49G>A variant in the UKBB cohort. The distribution of *FH* scores in the test cohorts is shown with solid violin plots, and 1000 Genomes controls with dashed violin plots. The 1000 Genomes critical values at the 99^th^ percentile are shown in horizontal lines. Samples confirmed to carry the variant based on WES analysis are shown in red. Samples without the variant that passed the 99^th^ percentile critical value for the Epi25 cohort are represented in black. The remaining samples are not represented individually. The critical values corresponding to the top 100 samples for the UKBB cohort are shown in brown horizontal lines for the UKBB cohort.

PR curve analysis wasn’t done for the Epi25 cohort due to the limited number of SCN1B carriers and the absence of WWOX carriers. In the UKBB cohort, the AUPRC of a random classifier is 0.00037 (carriers/total cohort size) for both variants ^54,55^, whereas FoundHaplo achieved an AUPRC of 0.46 for the *SCN1B* c.363C>G variant and 0.6 for the *WWOX* c.49G>A variant, indicating its effectiveness in distinguishing carriers from non-carriers in the UKBB cohort.

The total number of predictions above the 99^th^ percentile for the UKBB (n=9,523 for SCN1B c.363C>G and n=4,459 for *WWOX* c.49G>A) is typically too high for further screening (Tables S4-S5). For large cohorts, FoundHaplo can prioritize predictions by setting the selection of a specific number of samples with the highest *FH* scores for screening. Using this approach for the UKBB cohort, we assessed the top 100 samples for each of the two variants. This resulted in correctly predicting 53 carriers for the *SCN1B* c.363C>G variant and 74 carriers for the *WWOX* c.49G>A variant, with 31% and 43% sensitivity and with 53% and 74% true discovery rate, respectively (Tables S4-S5).

### Comparing FoundHaplo performance with DRIVE

We evaluated the ability of DRIVE ^23^ to identify the *SCN1B* c.363C>G and *WWOX* c.49G>A variant carriers in the Epi25 cohort. Carriers of each variant in the Epi25 cohort were inferred in two separate analyses by merging the Epi25 cohort with the original variant carriers that were used to define the disease haplotypes in FoundHaplo. DRIVE estimated two clusters at the *SCN1B* c.363C>G locus and seven clusters at the *WWOX* c.49G>A locus.

All five original *SCN1B* c.363C>G carriers clustered together in a single cluster. The two candidate *SCN1B* c.363C>G carriers in the Epi25 cohort were not clustered with any of the five original carriers. The three original *WWOX* c.49G>A carriers were clustered together. None of the other Epi25 samples were clustered with the original *SCN1B* c.363C>G or *WWOX* c.49G>A carriers. We could not evaluate the performance of DRIVE on the UKBB cohort as the sample size was too large, and the required run time was not feasible.

Based on the analysis, DRIVE did not detect the two *SCN1B* c.363C>G variant carriers in the Epi25, whereas FoundHaplo identified them at the 99^th^ percentile. However, DRIVE did not give any false positives for the two variants as none of the Epi25 samples clustered with any of the original variant carriers. FoundHaplo gave 13 and 23 false positives for the *SCN1B* c.363C>G and *WWOX* c.49G>A variants, respectively. However, the FoundHaplo analysis used an empirical 99% threshold; therefore, a 1% false positive rate is expected.

### Comparing FoundHaplo performance with genome-wide imputation tools

The *SCN1B* c.363C>G (p.Cys121Trp) and *WWOX* c.49G>A (p.E17K) variants were neither genotyped on SNP arrays, nor imputed by the MIS ^24^ or the TOPMed server ^25^ in the Epi25 cohort, possibly due to the absence or scarcity in variant-carrying haplotypes in reference panels. In the publicly available UKBB cohort, the *SCN1B* c.363C>G variant was imputed as heterozygous carriers in only nine samples out of 171 carriers (5% sensitivity) using the impute2 tool ^56^ with the HRC ^57^, UK10K ^58^ and 1000 Genomes phase 3 ^37^ reference panels. The *WWOX* c.49G>A variant is not imputed in the UKBB cohort, likely due to there being no *WWOX* c.49G>A carriers in the reference panel used in imputation. In contrast, FoundHaplo was able to correctly predict 55 *SCN1B* c.363C>G carriers and 74 *WWOX* c.49G>A carriers, using the 99^th^ critical percentile for the Epi25 and the top 100 samples for the UKBB cohort, showing a notable 37% sensitivity for both variants, which is a substantial improvement over genome-wide imputation tools.

## DISCUSSION

Achieving a genetic diagnosis is critical, providing opportunities for improved patient care by tailoring therapy appropriately, and potentially impacting the diagnosis of other family members, including distant relatives, who may also be at risk ^1,2^. With declining cost and widespread use of SNP genotyping, there are SNP arrays for millions of individuals in public databases. Hidden in these data are individuals who have inherited known, rare DCVs that are not ascertained directly by the SNP array and cannot be imputed due to their rarity.

FoundHaplo, uses SNP genotyping array data to identify individuals with rare DCVs based on known disease haplotypes, unlike traditional IBD algorithms that target genome-wide IBD regions ^16–22^. In our simulation study, FoundHaplo successfully detected 75% of cases sharing at least 2 cM of a disease haplotype. We evaluated the ability of FoundHaplo to identify two rare variants *SCN1B* c.363C>G (p.Cys121Trp) and *WWOX* c.49G>A (p.E17K), that can cause epilepsy: one a dominant allele, and the other a recessive variant requiring the presence of a second allele to cause disease. We showed that FoundHaplo’s targeted approach achieved 37% sensitivity in detecting the two rare epilepsy variants compared to the 5% sensitivity achieved using LD-based genomewide-imputation tools, despite the presence of multiple DCV-carrying haplotypes in the imputation reference panels, proving that FoundHaplo is superior to genome-wide imputation methods in inferring known DCVs using surrogate disease haplotypes. FoundHaplo also demonstrated greater sensitivity to DRIVE, a similar approach that was recently developed, however at the cost of a higher false positive rate.

We have shown that FoundHaplo can successfully identify disease haplotypes; however, the algorithm has a number of limitations. It uses a fixed error rate for genotype and imputation (1% by default), regardless of minor allele frequency (MAF) variations. A more refined method would adjust the error rate based on MAF., accommodating a higher error rate for rarer variants ^32,33,59,60^.

FoundHaplo doesn’t account for linkage disequilibrium (LD). While LD blocks are typically short and the algorithm targets longer IBD segments for the founder effects we seek to identify. incorporating LD might improve the detection of shorter IBD segments (≤1cM) with older common ancestors. The effect of LD can be seen in the simulation results, with performance varying by disease locus due to differences in background haplotype sharing, caused by locally specific LD, between controls.

FoundHaplo presumes accurate phasing of disease haplotypes, typically requiring multiple family members with a known DCV for pedigree phasing. Other LD-based genomewide phasing approaches, like those in TopMed or MIS, only offer block phasing. Additionally, FoundHaplo cannot determine the exact number of disease haplotype copies in a test individual. This is not relevant for autosomal dominant diseases since only one copy is sufficient to cause the disease. For recessive diseases, FoundHaplo can only predict individuals that carry at least one copy of the disease haplotype and further testing is required to determine the number of copies; however, this does not impact the utility of FoundHaplo as a screening tool. It will identify both carriers (one copy) and those individuals with two copies. These individuals may be homozygous or compound heterozygotes for inherited DCVs.

The power of the *FH* statistic increases the more unique disease haplotypes that are present. Additionally, FoundHaplo performs best when the disease and test individuals are more closely related to each other, allowing the preservation of a larger ancestral disease haplotype, and this is more likely to occur when there are more unique disease haplotypes present.

One important consideration when using FoundHaplo is the choice of critical threshold. The best choice depends on the appropriate balance between increasing sensitivity and minimising the number of false positives. The “false positives” identified by FoundHaplo that do not share the DCV may still share the disease haplotype since FoundHaplo uses disease haplotypes as surrogates for DCVs. This depends on the time between the DCV mutation and the uniqueness of the DCV-carrying haplotype prior to the DCV arising and is always unknown. For example, the *SCN1B* c.363C>G variant associated haplotype is present in ∼1% of the population ^6^, however, only a fraction of those individuals inherited the version of this haplotype with the DCV.

FoundHaplo, primarily designed for SNP genotyping data, can also utilize biallelic SNP genotypes from whole-genome sequencing (WGS) to identify disease haplotypes, which may be helpful for variants difficult to identify with short-read WGS, such as non-coding variants, cryptic splice variants and structural variants. We strongly recommend not using FoundHaplo on whole-exome sequencing (WES) data, where its effectiveness is likely limited due to sparse SNP markers, potentially increasing false negatives. FoundHaplo should be able to be used on any recombining genome for predicting inherited genetic variants.

The novelty of the FoundHaplo approach lies in using prior knowledge of known disease haplotypes to find local IBD segments specific to disease-causing variants of interest. We demonstrated the ability of FoundHaplo to detect two inherited rare variants that cause epilepsy. There are many other similar founder effects ideally suited for screening with this method. FoundHaplo could significantly aid in identifying carriers of known disease variants using SNP array data who might otherwise be unlikely to undergo targeted genetic testing or receive a genetic diagnosis.

## Data Availability

This research has been conducted using data from UK Biobank, a major biomedical database. The UK Biobank is an open access resource. To access the UKBB datasets, you need to register as a UKBB researcher (https://www.ukbiobank.ac.uk/enable-your-research/register). Additional genetic data used in this study is not available due to patient privacy and ethical restrictions.
FoundHaplo is available at https://github.com/bahlolab/FoundHaplo.

## Acknowledgements

We thank the Epi25 principal investigators, local staff from individual cohorts, and the individuals with epilepsy who participated in Epi25 for making possible this global collaboration and resource to advance epilepsy genetics research. This research was conducted with data from UK Biobank (www.ukbiobank.ac.uk), a major biomedical database, under data use agreement 36610 (PI Bahlo).

Funding support was provided by an Australian National Health and Medical Research Council (NHMRC) Investigator grant APP1195236 (MB); an NHMRC Senior Investigator Grant [GNT1172897] (IES); an NHMRC Senior Investigator Grant [APP196637] (SFB); a Melbourne Research Scholarship (392655) (ER); a CURE Epilepsy Taking Flight Award (MFB); an Australian Government Research Training Program Scholarship APP533086 (KLO); DHB Foundation Centenary Postdoctoral Fellowship in Neurogenetic Systems Biology (LGF); Health Research Council of New Zealand and Cure Kids (LS). This work was also supported by the Victorian Government’s Operational Infrastructure Support Program and the NHMRC Independent Research Institute Infrastructure Support Scheme. We would also like to acknowledge Professor Jozef Gecz and Dr Mark Corbett for valuable discussions on early work.

## Declaration of Interests

Ingrid Scheffer has served on scientific advisory boards for BioMarin, Chiesi, Eisai, Encoded Therapeutics, GlaxoSmithKline, Knopp Biosciences, Nutricia, Rogcon, Takeda Pharmaceuticals, UCB, Xenon Pharmaceuticals, Cerecin; has received speaker honoraria from GlaxoSmithKline, UCB, BioMarin, Biocodex, Chiesi, Liva Nova, Nutricia, Zuellig Pharma, Stoke Therapeutics and Eisai; has received funding for travel from UCB, Biocodex, GlaxoSmithKline, Biomarin, Encoded Therapeutics, Stoke Therapeutics and Eisai; has served as an investigator for Anavex Life Sciences, Cerevel Therapeutics, Eisai, Encoded Therapeutics, EpiMinder Inc, Epygenyx, ES-Therapeutics, GW Pharma, Marinus, Neurocrine BioSciences, Ovid Therapeutics, Takeda Pharmaceuticals, UCB, Ultragenyx, Xenon Pharmaceuticals, Zogenix and Zynerba; and has consulted for Care Beyond Diagnosis, Epilepsy Consortium, Atheneum Partners, Ovid Therapeutics, UCB, Zynerba Pharmaceuticals, BioMarin, Encoded Therapeutics and Biohaven Pharmaceuticals; and is a Non-Executive Director of Bellberry Ltd and a Director of the Australian Academy of Health and Medical Sciences and the Australian Council of Learned Academies Limited. She may accrue future revenue on pending patent WO61/010176 (filed: 2008): Therapeutic Compound; has a patent for *SCN1A* testing held by Bionomics Inc and licensed to various diagnostic companies; has a patent molecular diagnostic/theranostic target for benign familial infantile epilepsy (BFIE) [PRRT2] 2011904493 & 2012900190 and PCT/AU2012/001321 (TECH ID:2012-009).

## Web Resources

BCFtools, https://samtools.github.io/bcftools/bcftools.html

DRIVE, https://drive-ibd.readthedocs.io/en/latest/index.html

FoundHaplo, https://github.com/bahlolab/FoundHaplo

Genotype Harmonizer, https://github.com/molgenis/systemsgenetics/wiki/Genotype-Harmonizer

gnomAD, https://gnomad.broadinstitute.org/

HapMap project, https://www.genome.gov/10001688/international-hapmap-project

Michigan Imputation Server, https://imputationserver.sph.umich.edu/

OMIM, http://www.omim.org/

Plink 1.9, https://www.cog-genomics.org/plink/1.9/

SHAPEIT4, https://odelaneau.github.io/shapeit4/

TOPMed Imputation Server, https://imputation.biodatacatalyst.nhlbi.nih.gov/

UK Biobank, https://www.ukbiobank.ac.uk/

VCFtools, https://vcftools.github.io/index.html

1000 Genomes Project, https://www.internationalgenome.org/

## Data and code availability

This research has been conducted using data from UK Biobank, a major biomedical database. The UK Biobank is an open access resource. To access the UKBB datasets, you need to register as a UKBB researcher (https://www.ukbiobank.ac.uk/enable-your-research/register). Additional genetic data used in this study is not available due to patient privacy and ethical restrictions.

FoundHaplo is available at https://github.com/bahlolab/FoundHaplo.

## Notes

### Author Declarations

This study was approved by the Austin Health Human Research Ethics Committee. Informed consent was obtained and archived from all participants or their legal guardian. Research was approved by the Human Research Ethics Committee at The Walter and Eliza Hall Institute of Medical Research.

## REFERENCES

1. Lalonde, E., Rentas, S., Lin, F., Dulik, M.C., Skraban, C.M., and Spinner, N.B. (2020). Genomic Diagnosis for Pediatric Disorders: Revolution and Evolution. Front Pediatr 8, 373.

2. Marian, A.J. (2020). Clinical Interpretation and Management of Genetic Variants. JACC Basic Transl Sci 5, 1029–1042.

3. Bennett, M.F., Oliver, K.L., Regan, B.M., Bellows, S.T., Schneider, A.L., Rafehi, H., Sikta, N., Crompton, D.E., Coleman, M., Hildebrand, M.S., et al. (2020). Familial adult myoclonic epilepsy type 1 SAMD12 TTTCA repeat expansion arose 17,000 years ago and is present in Sri Lankan and Indian families. Eur. J. Hum. Genet. 28, 973–978.

4. Rafehi, H., Szmulewicz, D.J., Bennett, M.F., Sobreira, N.L.M., Pope, K., Smith, K.R., Gillies, G., Diakumis, P., Dolzhenko, E., Eberle, M.A., et al. (2019). Bioinformatics-Based Identification of Expanded Repeats: A Non-reference Intronic Pentamer Expansion in RFC1 Causes CANVAS. Am. J. Hum. Genet. 105, 151–165.

5. Albrechtsen, A., Sand Korneliussen, T., Moltke, I., van Overseem Hansen, T., Nielsen, F.C., and Nielsen, R. (2009). Relatedness mapping and tracts of relatedness for genome-wide data in the presence of linkage disequilibrium. Genet. Epidemiol. 33, 266–274.

6. Grinton, B.E., Robertson, E., Fearnley, L.G., Scheffer, I.E., Marson, A.G., O’Brien, T.J., Pickrell, W.O., Rees, M.I., Sisodiya, S.M., Balding, D.J., et al. (2022). A founder event causing a dominant childhood epilepsy survives 800 years through weak selective pressure. Am. J. Hum. Genet. 109, 2080–2087.

7. Paradisi, I., Hernández, A., and Arias, S. (2008). Huntington disease mutation in Venezuela: age of onset, haplotype analyses and geographic aggregation. J. Hum. Genet. 53, 127–135.

8. Henden, L., Freytag, S., Afawi, Z., Baldassari, S., Berkovic, S.F., Bisulli, F., Canafoglia, L., Casari, G., Crompton, D.E., Depienne, C., et al. (2016). Identity by descent fine mapping of familial adult myoclonus epilepsy (FAME) to 2p11.2–2q11.2. Hum. Genet. 135, 1117–1125.

9. McPeek, M.S., and Strahs, A. (1999). Assessment of linkage disequilibrium by the decay of haplotype sharing, with application to fine-scale genetic mapping. Am. J. Hum. Genet. 65, 858–875.

10. Henden, L., Twine, N.A., Szul, P., McCann, E.P., Nicholson, G.A., Rowe, D.B., Kiernan, M.C., Bauer, D.C., Blair, I.P., and Williams, K.L. (2020). Identity by descent analysis identifies founder events and links SOD1 familial and sporadic ALS cases. Npj Genomic Medicine 5, 1–8.

11. Nakashima K., Watanabe Y., Kusumi M., Nanba E., Maeoka Y., Igo M., Irie H., Ishino H., Fujimoto A., and Kobayashi S. (1995). [Prevalence and founder effect of Huntington’s disease in the San-in area of Japan]. Rinsho Shinkeigaku 35, 1532– 1534.

12. García-Planells, J., Burguera, J.A., Solís, P., Millán, J.M., Ginestar, D., Palau, F., and Espinós, C. (2005). Ancient origin of the CAG expansion causing Huntington disease in a Spanish population. Hum. Mutat. 25, 453–459.

13. Demetriou, C.A., Heraclides, A., Salafori, C., Tanteles, G.A., Christodoulou, K., Christou, Y., and Zamba-Papanicolaou, E. (2018). Epidemiology of Huntington disease in Cyprus: A 20-year retrospective study. Clin. Genet. 93, 656–664.

14. Morral, N., Bertranpetit, J., Estivill, X., Nunes, V., Casals, T., Giménez, J., Reis, A., Varon-Mateeva, R., Macek, M., Jr, and Kalaydjieva, L. (1994). The origin of the major cystic fibrosis mutation (delta F508) in European populations. Nat. Genet. 7, 169–175.

15. Boissé Lomax, L., Bayly, M.A., Hjalgrim, H., Møller, R.S., Vlaar, A.M., Aaberg, K.M., Marquardt, I., Gandolfo, L.C., Willemsen, M., Kamsteeg, E.-J., et al. (2013). “North Sea” progressive myoclonus epilepsy: phenotype of subjects with GOSR2 mutation. Brain 136, 1146–1154.

16. Naseri, A., Liu, X., Tang, K., Zhang, S., and Zhi, D. (2019). RaPID: ultra-fast, powerful, and accurate detection of segments identical by descent (IBD) in biobank-scale cohorts. Genome Biol. 20, 143.

17. Nait Saada, J., Kalantzis, G., Shyr, D., Cooper, F., Robinson, M., Gusev, A., and Palamara, P.F. (2020). Identity-by-descent detection across 487,409 British samples reveals fine scale population structure and ultra-rare variant associations. Nat. Commun. 11, 1–15.

18. Browning, B.L., and Browning, S.R. (2013). Improving the accuracy and efficiency of identity-by-descent detection in population data. Genetics 194, 459–471.

19. Gusev, A., Lowe, J.K., Stoffel, M., Daly, M.J., Altshuler, D., Breslow, J.L., Friedman, J.M., and Pe’er, I. (2009). Whole population, genome-wide mapping of hidden relatedness. Genome Res. 19, 318–326.

20. Purcell, S., Neale, B., Todd-Brown, K., Thomas, L., Ferreira, M.A.R., Bender, D., Maller, J., Sklar, P., de Bakker, P.I.W., Daly, M.J., et al. (2007). PLINK: a tool set for whole-genome association and population-based linkage analyses. Am. J. Hum. Genet. 81, 559–575.

21. Zhou, Y., Browning, S.R., and Browning, B.L. (2020). A Fast and Simple Method for Detecting Identity-by-Descent Segments in Large-Scale Data. Am. J. Hum. Genet. 106, 426–437.

22. Henden, L., Wakeham, D., and Bahlo, M. (2016). XIBD: software for inferring pairwise identity by descent on the X chromosome. Bioinformatics 32, 2389–2391.

23. Lancaster, M.C., Chen, H.-H., Shoemaker, M.B., Fleming, M.R., Baker, J.T., Polikowsky, H.G., Samuels, D.C., Huff, C.D., Roden, D.M., and Below, J.E. (2023). Detection of distant familial relatedness in biobanks for identification of undiagnosed carriers of a Mendelian disease variant: application to Long QT syndrome. medRxiv.

24. Das, S., Forer, L., Schönherr, S., Sidore, C., Locke, A.E., Kwong, A., Vrieze, S.I., Chew, E.Y., Levy, S., McGue, M., et al. (2016). Next-generation genotype imputation service and methods. Nat. Genet. 48, 1284–1287.

25. Taliun, D., Harris, D.N., Kessler, M.D., Carlson, J., Szpiech, Z.A., Torres, R., Taliun, S.A.G., Corvelo, A., Gogarten, S.M., Kang, H.M., et al. (2021). Sequencing of 53,831 diverse genomes from the NHLBI TOPMed Program. Nature 590, 290–299.

26. Sudlow, C., Gallacher, J., Allen, N., Beral, V., Burton, P., Danesh, J., Downey, P., Elliott, P., Green, J., Landray, M., et al. (2015). UK biobank: an open access resource for identifying the causes of a wide range of complex diseases of middle and old age. PLoS Med. 12, e1001779.

27. Suratannon, N., van Wijck, R.T.A., Broer, L., Xue, L., van Meurs, J.B.J., Barendregt, B.H., van der Burg, M., Dik, W.A., Chatchatee, P., Langerak, A.W., et al. (2020). Rapid Low-Cost Microarray-Based Genotyping for Genetic Screening in Primary Immunodeficiency. Front. Immunol. 11, 614.

28. Lu, C., Greshake Tzovaras, B., and Gough, J. (2021). A survey of direct-to-consumer genotype data, and quality control tool (GenomePrep) for research. Comput. Struct. Biotechnol. J. 19, 3747–3754.

29. Weedon, M.N., Jackson, L., Harrison, J.W., Ruth, K.S., Tyrrell, J., Hattersley, A.T., and Wright, C.F. (2019). Very rare pathogenic genetic variants detected by SNP-chips are usually false positives: implications for direct-to-consumer genetic testing.

30. Weedon, M.N., Jackson, L., Harrison, J.W., Ruth, K.S., Tyrrell, J., Hattersley, A.T., and Wright, C.F. (2021). Use of SNP chips to detect rare pathogenic variants: retrospective, population based diagnostic evaluation. BMJ 372,.

31. Auer, P.L., and Lettre, G. (2015). Rare variant association studies: considerations, challenges and opportunities. Genome Med. 7, 16.

32. Shi, S., Yuan, N., Yang, M., Du, Z., Wang, J., Sheng, X., Wu, J., and Xiao, J. (2018). Comprehensive Assessment of Genotype Imputation Performance. Hum. Hered. 83, 107–116.

33. De Marino, A., Mahmoud, A.A., Bose, M., Bircan, K.O., Terpolovsky, A., Bamunusinghe, V., Bohn, S., Khan, U., Novković, B., and Yazdi, P.G. (2022). A comparative analysis of current phasing and imputation software. PLoS One 17, e0260177.

34. Montgomery, G.W., Campbell, M.J., Dickson, P., Herbert, S., Siemering, K., Ewen-White, K.R., Visscher, P.M., and Martin, N.G. (2005). Estimation of the rate of SNP genotyping errors from DNA extracted from different tissues. Twin Res. Hum. Genet. 8, 346–352.

35. Stahl, K., Gola, D., and König, I.R. (2021). Assessment of Imputation Quality: Comparison of Phasing and Imputation Algorithms in Real Data. Front. Genet. 12, 724037.

36. Wilks, S.S. (1938). The Large-Sample Distribution of the Likelihood Ratio for Testing Composite Hypotheses. Ann. Math. Stat. 9, 60–62.

37. 1000 Genomes Project Consortium, Auton, A., Brooks, L.D., Durbin, R.M., Garrison, E.P., Kang, H.M., Korbel, J.O., Marchini, J.L., McCarthy, S., McVean, G.A., et al. (2015). A global reference for human genetic variation. Nature 526, 68–74.

38. International HapMap Consortium (2003). The International HapMap Project. Nature 426, 789–796.

39. International HapMap Consortium, Frazer, K.A., Ballinger, D.G., Cox, D.R., Hinds, D.A., Stuve, L.L., Gibbs, R.A., Belmont, J.W., Boudreau, A., Hardenbol, P., et al. (2007). A second generation human haplotype map of over 3.1 million SNPs. Nature 449, 851–861.

40. Karczewski, K.J., Francioli, L.C., Tiao, G., Cummings, B.B., Alföldi, J., Wang, Q., Collins, R.L., Laricchia, K.M., Ganna, A., Birnbaum, D.P., et al. (2020). The mutational constraint spectrum quantified from variation in 141,456 humans. Nature 581, 434– 443.

41. Chen, S., Francioli, L.C., Goodrich, J.K., Collins, R.L., Kanai, M., Wang, Q., Alföldi, J., Watts, N.A., Vittal, C., Gauthier, L.D., et al. (2023). A genomic mutational constraint map using variation in 76,156 human genomes. Nature.

42. Li, Y., Willer, C.J., Ding, J., Scheet, P., and Abecasis, G.R. (2010). MaCH: using sequence and genotype data to estimate haplotypes and unobserved genotypes. Genet. Epidemiol. 34, 816–834.

43. Howie, B., Fuchsberger, C., Stephens, M., Marchini, J., and Abecasis, G.R. (2012). Fast and accurate genotype imputation in genome-wide association studies through pre-phasing. Nat. Genet. 44, 955–959.

44. Depienne, C., and Mandel, J.-L. (2021). 30 years of repeat expansion disorders: What have we learned and what are the remaining challenges? Am. J. Hum. Genet. 108, 764–785.

45. Browning, B.L., and Browning, S.R. (2022). Genotype error biases trio-based estimates of haplotype phase accuracy. Am. J. Hum. Genet. 109, 1016–1025.

46. Zhang, Y.-H., Burgess, R., Malone, J.P., Glubb, G.C., Helbig, K.L., Vadlamudi, L., Kivity, S., Afawi, Z., Bleasel, A., Grattan-Smith, P., et al. (2017). Genetic epilepsy with febrile seizures plus: Refining the spectrum. Neurology 89, 1210–1219.

47. Wallace, R.H., Scheffer, I.E., Parasivam, G., Barnett, S., Wallace, G.B., Sutherland, G.R., Berkovic, S.F., and Mulley, J.C. (2002). Generalized epilepsy with febrile seizures plus: mutation of the sodium channel subunit SCN1B. Neurology 58, 1426–1429.

48. Scheffer, I.E., Harkin, L.A., Grinton, B.E., Dibbens, L.M., Turner, S.J., Zielinski, M.A., Xu, R., Jackson, G., Adams, J., Connellan, M., et al. (2007). Temporal lobe epilepsy and GEFS+ phenotypes associated with SCN1B mutations. Brain 130, 100– 109.

49. Patino, G.A., Claes, L.R.F., Lopez-Santiago, L.F., Slat, E.A., Dondeti, R.S.R., Chen, C., O’Malley, H.A., Gray, C.B.B., Miyazaki, H., Nukina, N., et al. (2009). A functional null mutation of SCN1B in a patient with Dravet syndrome. J. Neurosci. 29, 10764–10778.

50. Abdel-Salam, G., Thoenes, M., Afifi, H.H., Körber, F., Swan, D., and Bolz, H.J. (2014). The supposed tumor suppressor gene WWOX is mutated in an early lethal microcephaly syndrome with epilepsy, growth retardation and retinal degeneration. Orphanet J. Rare Dis. 9, 12.

51. Mallaret, M., Synofzik, M., Lee, J., Sagum, C.A., Mahajnah, M., Sharkia, R., Drouot, N., Renaud, M., Klein, F.A.C., Anheim, M., et al. (2014). The tumour suppressor gene WWOX is mutated in autosomal recessive cerebellar ataxia with epilepsy and mental retardation. Brain 137, 411–419.

52. Oliver, K.L., Trivisano, M., Mandelstam, S.A., De Dominicis, A., Francis, D.I., Green, T.E., Muir, A.M., Chowdhary, A., Hertzberg, C., Goldhahn, K., et al. (2023). WWOX developmental and epileptic encephalopathy: Understanding the epileptology and the mortality risk. Epilepsia 64, 1351–1367.

53. Feng, Y.-C.A., Howrigan, D.P., Abbott, L.E., Tashman, K., Cerrato, F., Singh, T., Heyne, H., Byrnes, A., Churchhouse, C., Watts, N., et al. (2019). Ultra-Rare Genetic Variation in the Epilepsies: A Whole-Exome Sequencing Study of 17,606 Individuals. Am. J. Hum. Genet. 105, 267–282.

54. Miao, J., and Zhu, W. (2022). Precision–recall curve (PRC) classification trees. Evol. Intell. 15, 1545–1569.

55. Saito, T., and Rehmsmeier, M. (2015). The precision-recall plot is more informative than the ROC plot when evaluating binary classifiers on imbalanced datasets. PLoS One 10, e0118432.

56. Howie, B.N., Donnelly, P., and Marchini, J. (2009). A flexible and accurate genotype imputation method for the next generation of genome-wide association studies. PLoS Genet. 5, e1000529.

57. McCarthy, S., Das, S., Kretzschmar, W., Delaneau, O., Wood, A.R., Teumer, A., Kang, H.M., Fuchsberger, C., Danecek, P., Sharp, K., et al. (2016). A reference panel of 64,976 haplotypes for genotype imputation. Nat. Genet. 48, 1279–1283.

58. UK10K Consortium, Walter, K., Min, J.L., Huang, J., Crooks, L., Memari, Y., McCarthy, S., Perry, J.R.B., Xu, C., Futema, M., et al. (2015). The UK10K project identifies rare variants in health and disease. Nature 526, 82–90.

59. Ausmees, K., and Nettelblad, C. (2023). Achieving improved accuracy for imputation of ancient DNA. Bioinformatics 39,.

60. Phocas, F. (2022). Genotyping, the Usefulness of Imputation to Increase SNP Density, and Imputation Methods and Tools. Methods Mol. Biol. 2467, 113–138.

