## Supplemental Information for "Identifying individuals with rare disease variants by inferring shared ancestral haplotypes from SNP array data"

### Supplemental Figures and Tables

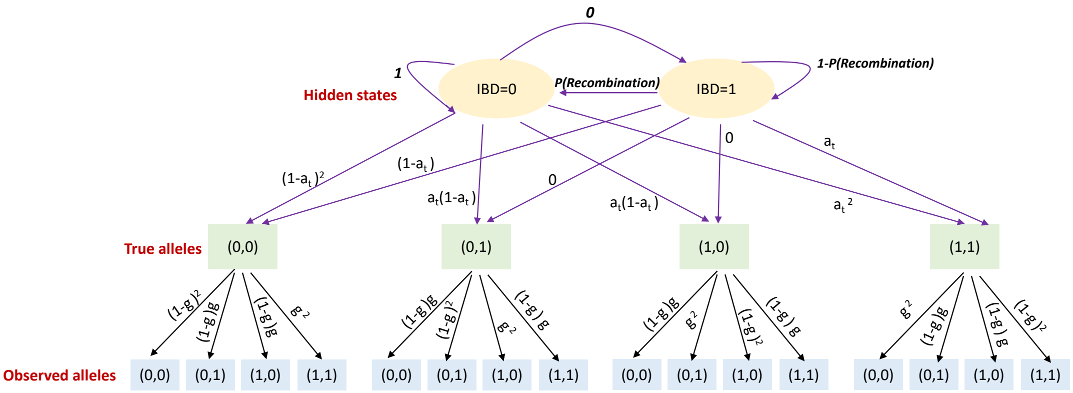


**Figure S1:** Transition and emission probabilities of the FoundHaplo Hidden Markov model. The genotype and imputation error rate is denoted by $g$ and the minor allele frequency by $a$.

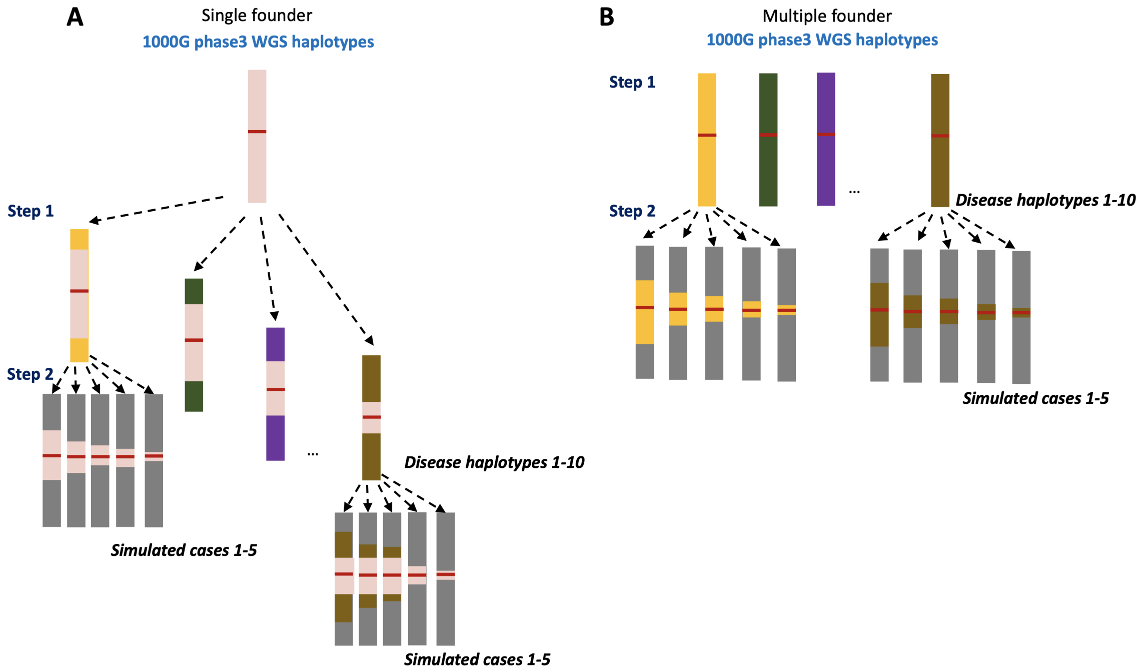


**Figure S2:** Simulating IBD segments for single and multiple founder effects. Simulating IBD segments between individuals with two models of founder effects, (A) single founder effect simulations and (B) multiple founder effect simulations. The disease variant is shown in a red horizontal line. Two steps are involved in simulating IBD segments in single founder effect models. Step 1: Randomly select disease haplotypes and simulate them to inherit the core IBD haplotype from their common ancestor. Step 2: Simulate IBD segments in chosen cases to inherit the chosen disease haplotypes. Both steps 1 and 2 are performed by replacing varying in-size genomic regions around the disease variant of the predecessors to their respective successors. Step 2 is similar in both single and multiple founder effects. In multiple founder effect simulations during step 1, we randomly select unique disease haplotypes, i.e. there are multiple common ancestors. The expected values of the total size of the replaced haplotypes in step 1 are set as 1, 2, 3, 4, 5, 6, 7, 8, 9 and 10 cM, reflecting 200 generations to 20 generations since the common ancestor. The expected values of the total size of the replaced haplotypes in step 2 are chosen to be 0.5, 1, 2 and 5 cM, reflecting 400 generations to 40 generations since the common ancestor. The distances in cM from the DCV locus to the left or right breakpoints of the ancestral segments in step 1 and step 2 were simulated using independent exponential random variables [1].

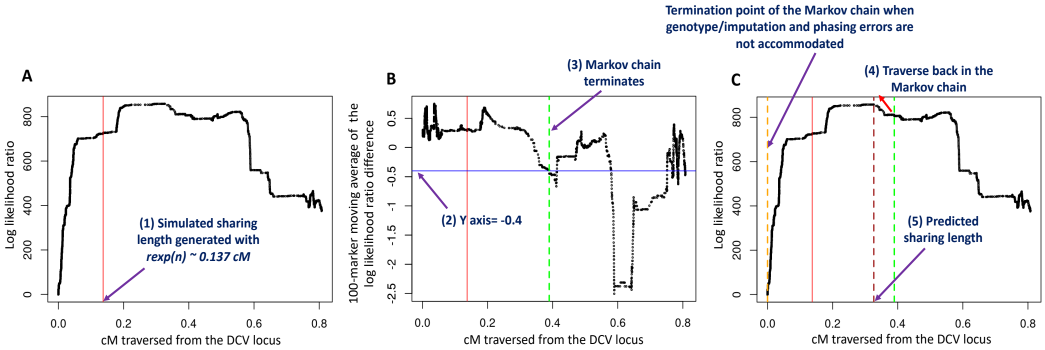


**Figure S3:** Deriving the stopping criteria in the Markov chains based on the simulation results. (A) Change of log-likelihood ratio (*LLR*) in the Markov chain. The IBD sharing endpoint is denoted by a red vertical line. The *LLR* starts to gradually decrease after the IBD sharing endpoint. (B) Based on the simulation results, the Markov chain is terminated when the 100-marker moving average (calculated by taking 100 adjacent SNP markers) of the difference between adjacent *LLR* values decreases below -0.4, which is shown in a green vertical line. (C) Once the Markov chain is terminated, the algorithm traverses backwards in the chain to identify the point at which the Markov chain gave the highest *LLR* score, which is shown in a dashed vertical red line and is taken as the termination point in the Markov chain. The orange vertical line shows the point at which the Markov chain would have terminated if genotype/imputation and phasing errors were not accommodated. Results are shown only for a single simulation.

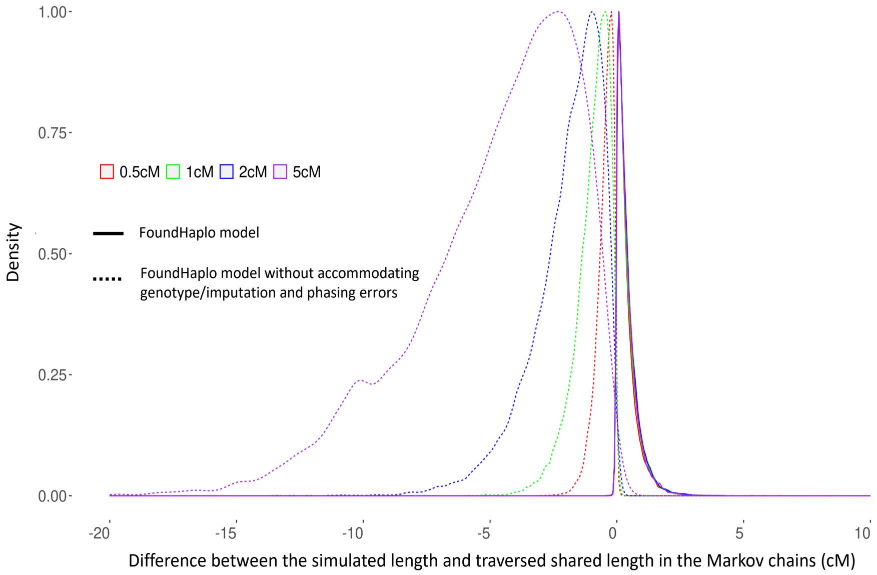


**Figure S4:** Density of the difference between the predicted and the expected length of haplotype sharing of simulated cases based on the simulation results. The FoundHaplo algorithm terminated before the simulated sharing length when the genotype/imputation and phasing errors were not accommodated.

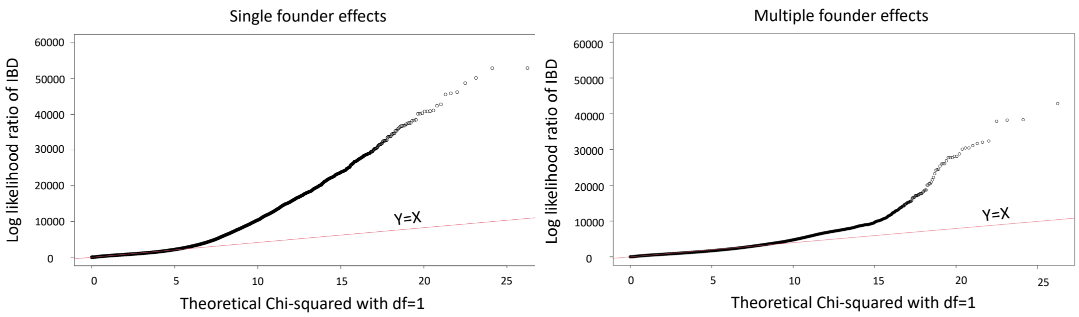


**Figure S5:** Q-Q plots comparing the log-likelihood ratio values. Q-Q plots comparing the log-likelihood ratio values under the null hypothesis (Y-axis) against the quantiles of the chi-squared distribution with one degree of freedom. Y-axis represents the log-likelihood ratio (*LLR*) values for simulated controls across all simulations (33 DCVs × 10 founders × 442 controls= 145,860 data points), which follow the null hypothesis. The Y=X line is shown in red. The *LLR* values under the null hypothesis deviate from the chi-squared distribution with one degree of freedom.

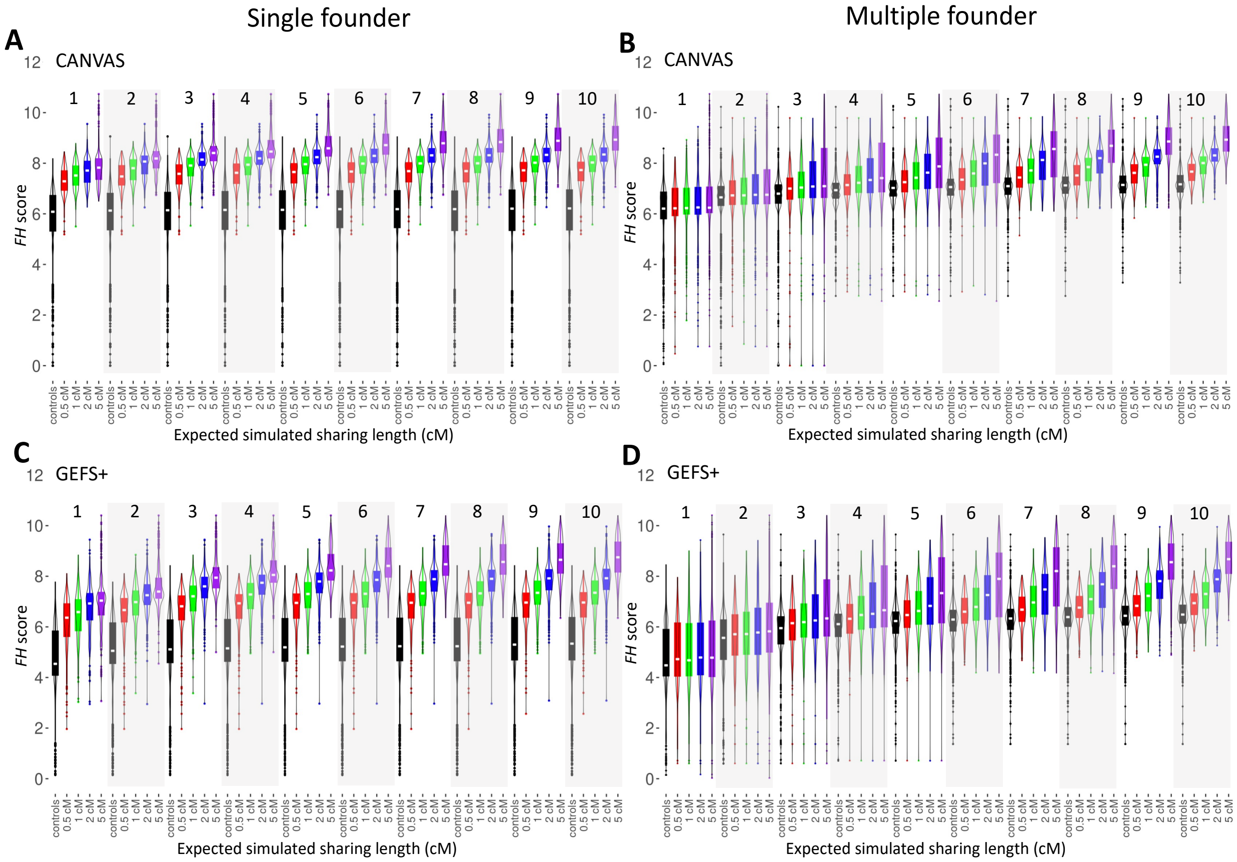


**Figure S6:** *FH* scores of simulated cases and controls for CANVAS and GEFS+ loci. *FH* scores of simulated cases: 0.5 cM (red), 1 cM (green), 2 cM (blue) and 5 cM (purple), and controls (black) for (A) CANVAS locus for single founder effects, (B) CANVAS locus for multiple founder effects, (C) GEFS+ locus for single founder effects and (D) GEFS+ locus for multiple founder effects. Results for simulations that include increasing numbers of disease haplotypes are partitioned for better visualisation.

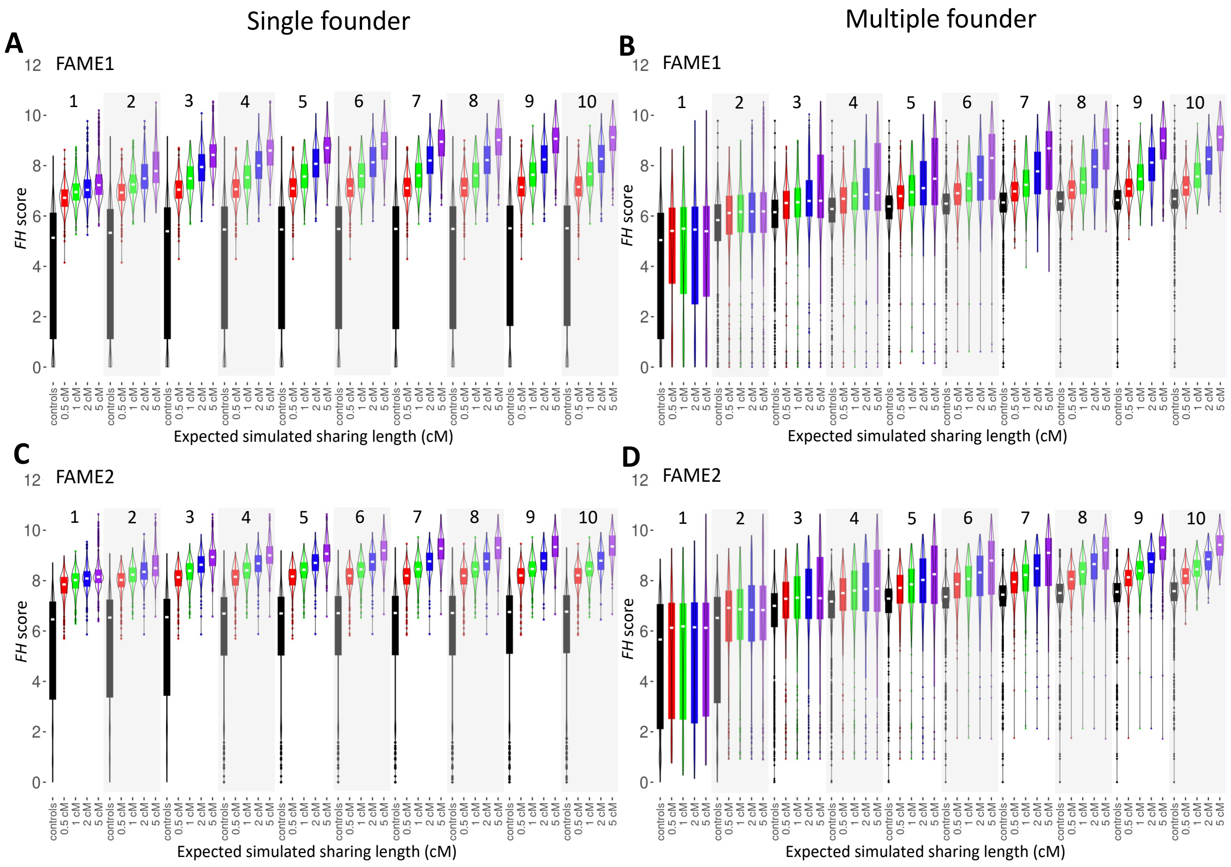


**Figure S7:** *FH* scores of simulated cases and controls for FAME1 and FAME2 loci. *FH* scores of simulated cases: 0.5 cM (red), 1 cM (green), 2 cM (blue) and 5 cM (purple), and controls (black) for (A) FAME1 locus for single founder effects, (B) FAME1 locus for multiple founder effects, (C) FAME2 locus for single founder effects and (D) FAME2 locus for multiple founder effects. Results for simulations that include increasing numbers of disease haplotypes are partitioned for better visualisation.

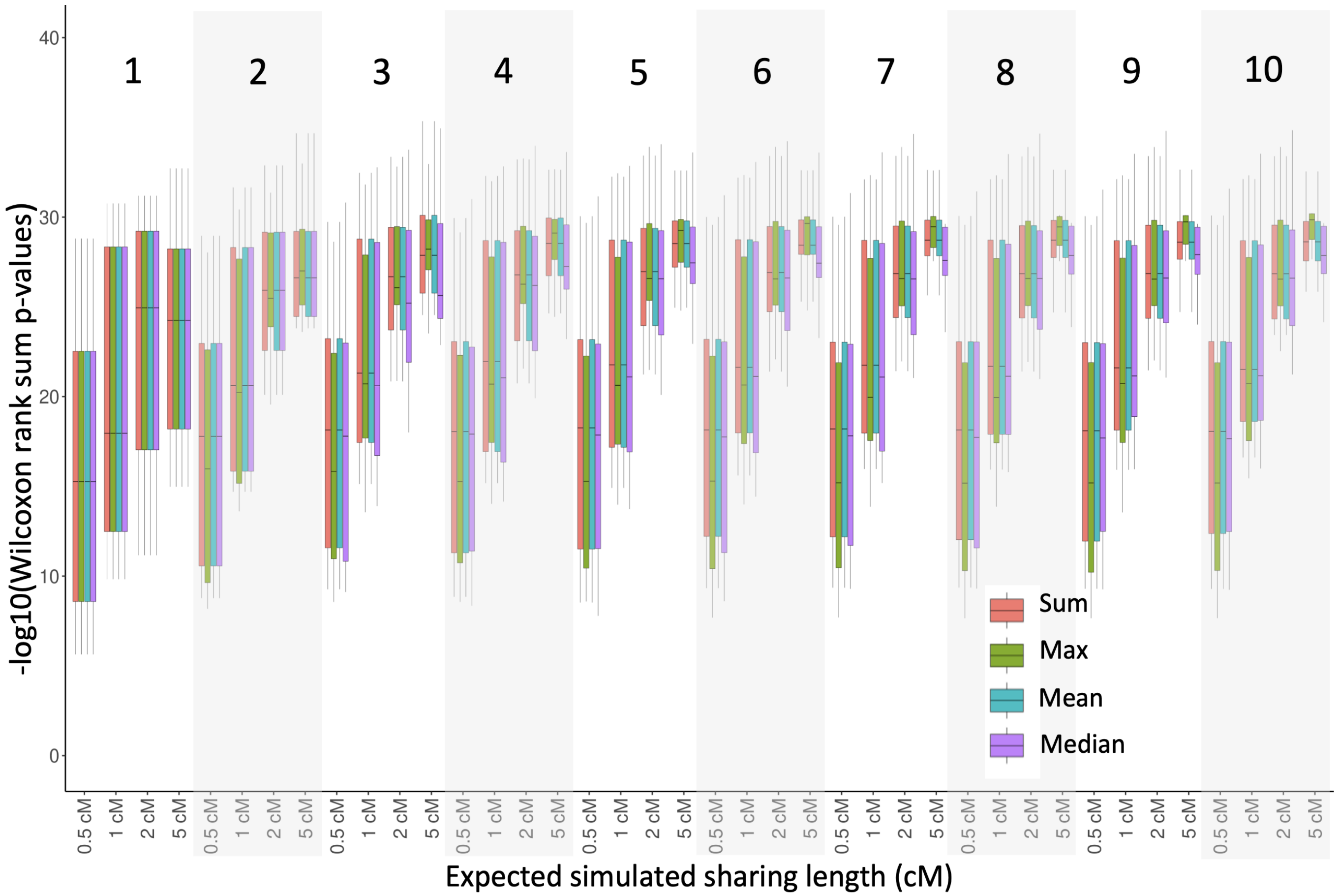


**Figure S8:** Comparing different approaches for combining multiple $FH$ scores at a locus. Box plots represent the Wilcoxon rank sum p-values for simulated cases versus controls when multiple disease haplotypes are present for single founder effect simulations averaged across all 33 DCVs. All four measures (summation($FH$), maximum($FH$), mean($FH$) and median($FH$)) performed similarly in single founder effect simulations. Results for the 1-10 number of disease haplotypes are partitioned for better visualisation.

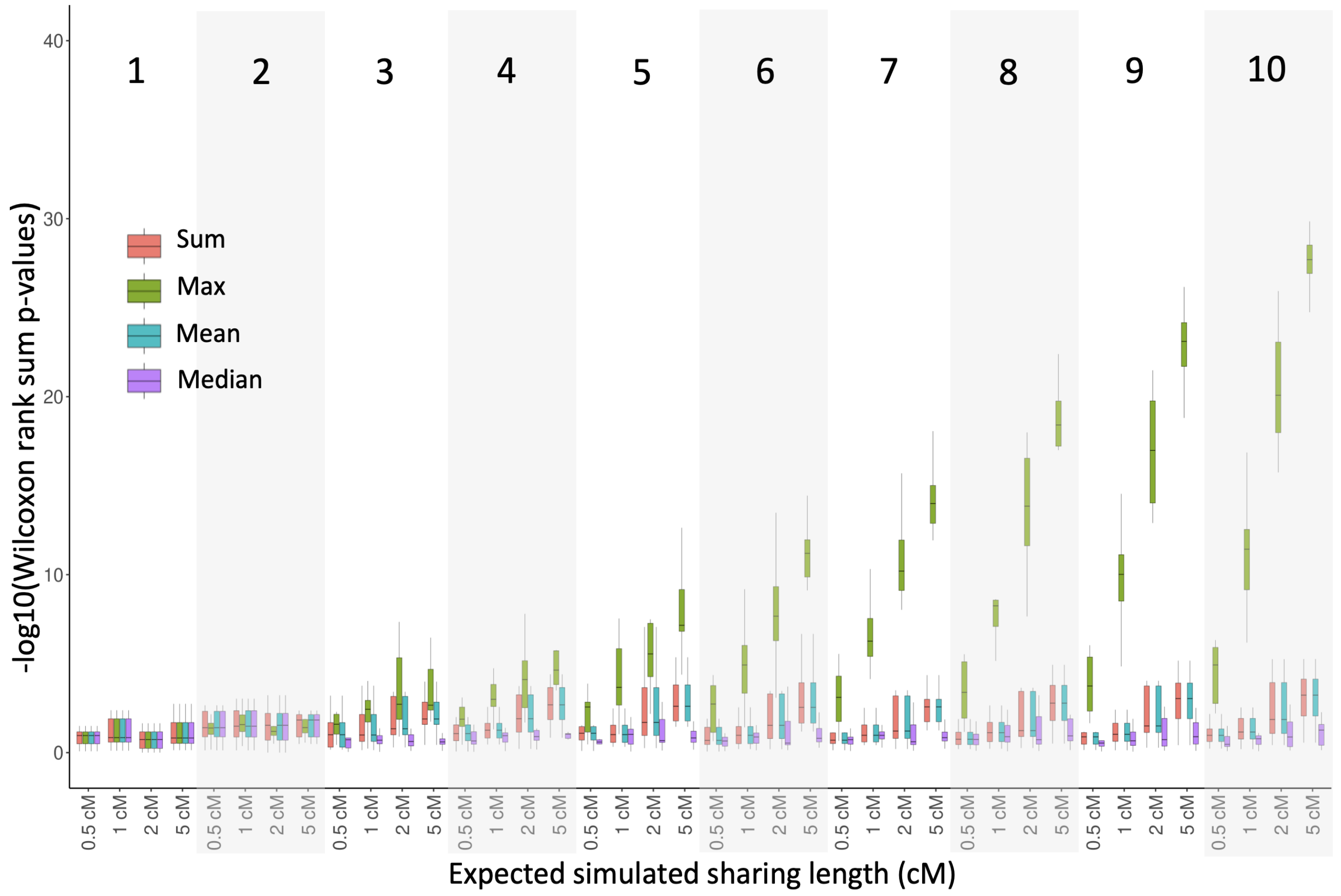


**Figure S9:** Comparing different approaches for combining multiple *FH* scores at a locus. Box plots represent the Wilcoxon rank sum p-values for simulated cases versus controls when multiple disease haplotypes are present for multiple founder effect simulations averaged across all 33 DCVs. The maximum of individual *FH* scores gave the highest -log10(P-value) based on the Wilcoxon rank sum test for multiple founder effect simulations. Results for the 1-10 number of disease haplotypes are partitioned for better visualisation.

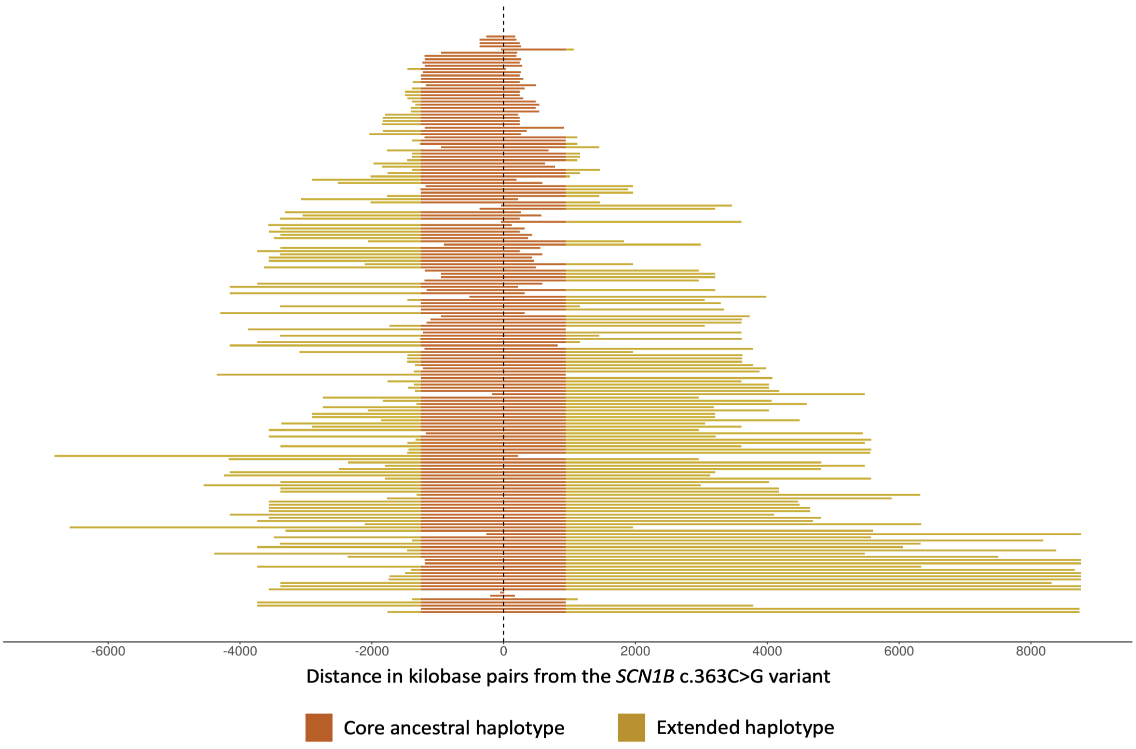


**Figure S10:** Shared haplotype regions identified in carriers of the *SCN1B* c.363C>G variant. Shared haplotype regions identified in the five *SCN1B* c.363C>G disease haplotypes, two Epi25 samples and the 171 UKBB samples known to carry the *SCN1B* c.363C>G variant (in the order of largest to smallest shared regions in each cohort from bottom to top). The location of the *SCN1B* c.363C>G variant is shown by the dotted line. Dark orange represents the 4.1 cM core ancestral region shared by all the five disease haplotypes. Yellow represents the regions shared with at least one of the five disease haplotypes. Both Epi25 samples and 82 UKBB samples did not share the entire core haplotype. All the samples shared a haplotype of at least 55 kbps. Only the markers with imputation accuracy 0.9 were retained when evaluating the haplotype sharing.

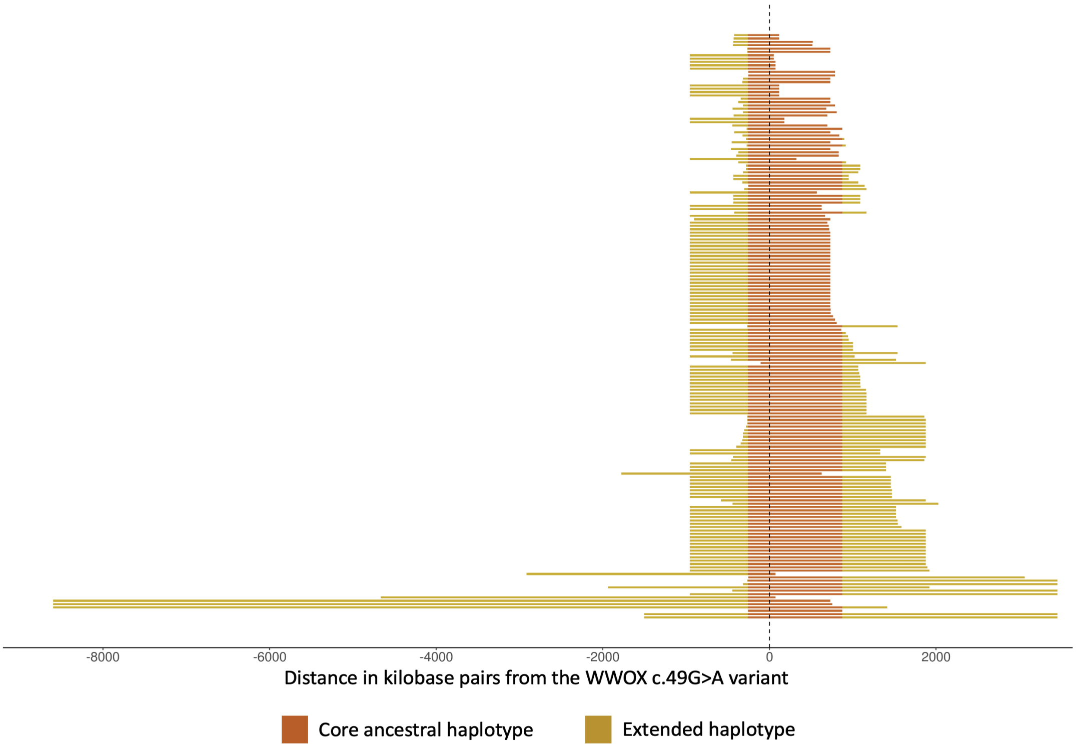


**Figure S11:** Shared haplotype regions identified in carriers of the *WWOX* c.49G>A varint. Shared haplotype regions identified in the three *WWOX* c.49G>A disease haplotypes and the 172 UKBB samples known to carry the *WWOX* c.49G>A variant (in the order of largest to smallest shared regions in each cohort from bottom to top). The location of the *WWOX* c.49G>A variant is shown by the dotted line. Dark orange represents the 3.9 cM core ancestral region shared by all three disease haplotypes. Yellow represents the regions shared with at least one of the three disease haplotypes. Eighty-one UKBB samples did not share the entire core haplotype. All the samples shared a haplotype of at least 157 kbps. Only the markers with imputation accuracy 0.9 were retained when evaluating the haplotype sharing.

| Associated disease | OMIM abbreviation | OMIM ID | Disease-causing variant | Inheritence | Gene | Genomic region | Founder origin |
|---|---|---|---|---|---|---|---|
| Cerebellar ataxia, neuropathy, vestibular areflexia syndrome | CANVAS | 614575 | TTCCC repeat expansion | AR | *RFC1* | intron | Europe [2,3] |
| Ceroid lipofuscinosis, neuronal, 3 | CLN3 | 204200 | *CLN3* c.461-280_677+382del966 | AR | *CLN3* | intron | Europe [4] |
| Cystic Fibrosis | CF | 219700 | *CFTR* p.F508del | AR | *CFTR* | coding | Europe [5] |
| Dentatorubral-pallidoluysian atrophy | DRPLA | 125370 | CAG repeat expansion | AD | *ATN1* | coding | Portugal [6], Italy [7] |
| Developmental and epileptic encephalopathy 28 | DEE28 | 616211 | *WWOX* c.49G>A | AR | *WWOX* | coding | Yemenite Jews [8] |
| Familial adult myoclonic epilepsy type 1 | FAME1 | 601068 | TTTCA repeat expansion | AD | *SAMD12* | intron | Asia [9–12] |
| Familial adult myoclonic epilepsy type 2 | FAME2 | 607876 | TTTCA repeat expansion | AD | *STARD7* | intron | Italy [13,14] |
| Familial adult myoclonic epilepsy type 3 | FAME3 | 613608 | TTTCA repeat expansion | AD | *MARCHF6* | intron | Europe [15] |
| Fragile X syndrome | FRAXA | 300624 | CGG repeat expansion | XLD | *FMR1* | 5'UTR | Cameroon [16], Finland [17], Sweden [18] |
| Friedreich ataxia | FRDA | 229300 | GAA repeat expansion | AR | *FXN* | intron | France [19], Spain [20], Louisiana Acadians [21], Germany [22] |
| Frontotemporal dementia and/or amyotrophic lateral sclerosis 1 | FTDALS1 | 105550 | GGGGCC repeat expansion | AD | *C9orf72* | intron | Europe [23], Finland [24,25], Japan [26] |

| Associated disease | OMIM abbreviation | OMIM ID | Disease-causing variant | Inheritence | Gene | Genomic region | Founder origin |
|---|---|---|---|---|---|---|---|
| Genetic epilepsy with febrile seizures plus, type 1 | GEFS+ | 604233 | *SCN1B c.363C>G* | AD | *SCN1B* | coding | Europe [27] |
| Huntington's disease | HD | 143100 | CAG repeat expansion | AD | *HTT* | coding | Venezuela [28], Japan [29], Spain [30], Europe [31] |
| Myotonic dystrophy 1 | DM1 | 160900 | CTG repeat expansion | AD | *DMPK* | 3'UTR | Northeastern Quebec [32], South Africa [33] |
| Myoclonic epilepsy of Unverricht and Lundborg | EPM1 | 254800 | CCCCGCCCCGCG repeat expansion | AR | *CSTB* | promotor | Reunion Island, a French island in the Indian Ocean [34] |
| Progressive myoclonic epilepsy | EPM6 | 614018 | *GOSR2 c.430G>T* | AR | *GOSR2* | Coding | Europe [35] |
| Spinal and bulbar muscular atrophy of Kennedy | SBMA | 313200 | CAG repeat expansion | XLR | *AR* | coding | Japan [36], Danish [37], Scandinavia [38] |
| Spinocerebellar ataxia 2 | SCA2 | 183090 | CAG repeat expansion | AD | *ATXN2* | coding | Cuba [39], Japan [40] |
| Spinocerebellar ataxia 3 | SCA3 | 109150 | CAG repeat expansion | AD | *ATXN3* | coding | Common founder for French,Portuguese-Brazilian,Portuguese-Azorean [41], China [42] |
| Spinocerebellar ataxia 6 | SCA6 | 183086 | CAG repeat expansion | AD | *CACNA1A* | coding | Japan [43], Germany [44] |
| Spinocerebellar ataxia 10 | SCA10 | 603516 | ATTCT repeat expansion | AD | *ATXN10* | intron | Latin America [45], Native American [46] |
| Spinocerebellar ataxia 12 | SCA12 | 604326 | CAG repeat expansion | AD | *PPP2R2B* | promotor | India [47] |
| Spinocerebellar ataxia 17 | SCA17 | 607136 | CAG repeat expansion | AD | *TBP* | coding | Germany [48] |
| Spinocerebellar ataxia 31 | SCA31 | 117210 | TGGAA repeat expansion | AD | *BEAN1* | intron | Japan [49–51] |

AD: Autosomal dominant inheritance, AR: Autosomal recessive inheritance, XLD: X-linked dominant inheritance, and XLR: X-linked recessive inheritance.

**Table S1:** Disease-causing variants with known founder effects.

| $o_t^x$ | $o_t^{y_h}$ | $o\prime_t^x$ | $o\prime_t^{y_h}$ | $P(o_t^{x,y_h}, o\prime_t^{x,y_h} \mid q_t^{x,y_h} = 0)$ | $P(o_t^{x,y_h}, o\prime_t^{x,y_h} \mid q_t^{x,y_h} = 1)$ |
|---|---|---|---|---|---|
| 0 | 0 | 0 | 0 | $((1-g)(1-a_t))^2$ | $(1-g)^2(1-a_t)$ |
| 0 | 0 | 0 | 1 | $(1-g)g(1-a_t)a_t$ | 0 |
| 0 | 0 | 1 | 0 | $(1-g)g(1-a_t)a_t$ | 0 |
| 0 | 0 | 1 | 1 | $(ga_t)^2$ | $g^2a_t$ |
| 0 | 1 | 0 | 0 | $(1-g)g(1-a_t)^2$ | $(1-g)g(1-a_t)$ |
| 0 | 1 | 0 | 1 | $(1-g)^2(1-a_t)a_t$ | 0 |
| 0 | 1 | 1 | 0 | $g^2(1-a_t)a_t$ | 0 |
| 0 | 1 | 1 | 1 | $(1-g)ga_t^2$ | $(1-g)ga_t$ |
| 1 | 0 | 0 | 0 | $(1-g)g(1-a_t)^2$ | $(1-g)g(1-a_t)$ |
| 1 | 0 | 0 | 1 | $g^2(1-a_t)a_t$ | 0 |
| 1 | 0 | 1 | 0 | $(1-g)^2(1-a_t)a_t$ | 0 |
| 1 | 0 | 1 | 1 | $(1-g)ga_t^2$ | $(1-g)ga_t$ |
| 1 | 1 | 0 | 0 | $(g(1-a_t))^2$ | $g^2(1-a_t)$ |
| 1 | 1 | 0 | 1 | $(1-g)g(1-a_t)a_t$ | 0 |
| 1 | 1 | 1 | 0 | $(1-g)g(1-a_t)a_t$ | 0 |
| 1 | 1 | 1 | 1 | $((1-g)a_t)^2$ | $(1-g)^2a_t$ |

**Table S2:** Emission probabilities $B$ in the FoundHaplo algorithm.

| **Associated disease** | **OMIM abbreviation** | **OMIM ID** | **Inheritance** | **Chromosome** | **hg19 base pair position** | **Gene** | **Genomic region** |
|---|---|---|---|---|---|---|---|
| Cerebellar ataxia, neuropathy, and vestibular areflexia syndrome | CANVAS | 614575 | AD | chr4 | 39350045 | *RFC1* | intron |
| Corneal dystrophy, Fuchs endothelial, 3 | FECD3 | 613267 | AD | chr18 | 53253385 | *TCF4* | intron |
| Dentatorubral-pallidoluysian atrophy | DRPLA | 125370 | AD | chr12 | 7045880 | *ATN1* | coding |
| Familial adult myoclonic epilepsy 1 | FAME1 | 601068 | AD | chr8 | 119379052 | *SAMD12* | intron |
| Familial adult myoclonic epilepsy 2 | FAME2 | 607876 | AD | chr2 | 96862805 | *STARD7* | intron |
| Familial adult myoclonic epilepsy 3 | FAME3 | 613608 | AD | chr5 | 10356520 | *MARCHF6* | intron |
| Familial adult myoclonic epilepsy 4 | FAME4 | 615127 | AD | chr3 | 183430011 | *YEATS2* | intron |
| Familial adult myoclonic epilepsy 6 | FAME6 | 618074 | AD | chr16 | 24624851 | *TNRC6A* | intron |
| Familial adult myoclonic epilepsy 7 | FAME7 | 618075 | AD | chr4 | 160263769 | *RAPGEF2* | intron |
| Fragile X syndrome | FRAXA | 300624 | XLD | chrX | 146993555 | *FMR1* | 5'UTR |
| Friedreich ataxia | FRDA | 229300 | AR | chr9 | 71652201 | *FXN* | intron |
| Genetic epilepsy with febrile seizures plus, type 1 | GEFS+ | 604233 | AD | chr19 | 35524558 | *SCN1B* | coding |
| Global developmental delay, progressive ataxia, and elevated glutamine | *GDPAG* | 618412 | AR | chr2 | 191745599 | *GLS* | 5'UTR |
| Huntington's disease | HD | 143100 | AD | chr4 | 3076604 | *HTT* | coding |
| Huntington disease-like 2 | HDL2 | 606438 | AD | chr16 | 87637889 | *JPH3* | exon |
| Intellectual developmen- tal disorder, X-linked 109 | FRAXE | 309548 | XLR | chrX | 147582159 | *AFF2* | 5'UTR |
| Myotonic dystrophy 1 | DM1 | 160900 | AD | chr19 | 46273463 | *DMPK* | 3'UTR |
| Myotonic dystrophy 2 | DM2 | 602668 | AD | chr3 | 128891420 | CNBP | intron |

| Associated disease | OMIM abbreviation | OMIM ID | Inheritance | Chromosome | Position | Gene | Genomic region |
|---|---|---|---|---|---|---|---|
| Myoclonic epilepsy of Unverricht and Lundborg | EPM1 | 254800 | AR | chr21 | 45196324 | *CSTB* | 5'UTR |
| Spinal and bulbar muscular atrophy of Kennedy | SBMA | 313200 | XLR | chrX | 66765159 | *AR* | coding |
| Spinocerebellar ataxia 1 | SCA1 | 164400 | AD | chr6 | 16327865 | ATXN1 | coding |
| Spinocerebellar ataxia 2 | SCA2 | 183090 | AD | chr12 | 112036754 | *ATXN2* | coding |
| Spinocerebellar ataxia 3 | SCA3 | 109150 | AD | chr14 | 92537355 | *ATXN3* | coding |
| Spinocerebellar ataxia 6 | SCA6 | 183086 | AD | chr19 | 13318673 | *CACNA1A* | coding |
| Spinocerebellar ataxia 7 | SCA7 | 164500 | AD | chr3 | 63898361 | *ATXN7* | coding |
| Spinocerebellar ataxia 8 | SCA8 | 608768 | AD | chr13 | 70713516 | *ATXN8OS* | utRNA |
| Spinocerebellar ataxia 10 | SCA10 | 603516 | AD | chr22 | 46191235 | *ATXN10* | intron |
| Spinocerebellar ataxia 12 | SCA12 | 604326 | AD | chr5 | 146258291 | *PPP2R2B* | promotor |
| Spinocerebellar ataxia 17 | SCA17 | 607136 | AD | chr6 | 170870995 | *TBP* | coding |
| Spinocerebellar ataxia 31 | SCA31 | 117210 | AD | chr16 | 066524302 | *BEAN1* | intron |
| Spinocerebellar ataxia 36 | SCA36 | 614153 | AD | chr20 | 2633379 | *NOP56* | intron |
| Spinocerebellar ataxia 37 | SCA37 | 615945 | AD | chr1 | 057832716 | *DAB1* | intron |

AD: Autosomal dominant inheritance, AR: Autosomal recessive inheritance, XLD: X-linked dominant inheritance, and XLR: X-linked recessive inheritance.

**Table S3:** List of disease loci used to evaluate the performance of FoundHaplo in the simulation study.

| Epi25 | | | | | | UKBB | | | | | |
|---|---|---|---|---|---|---|---|---|---|---|---|
| **Critical percentile (%)** | **Samples predicted above the critical value** | **TP % (number)** | **FP % (number)** | **TD % (number)** | **FD % (number)** | **Critical percentile (%)** | **Samples predicted above the critical value** | **TP % (number)** | **FP % (number)** | **TD % (number)** | **FD % (number)** |
| 99 | 15 | 100% (2) | 0.8% (13) | 13% (2) | 87% (13) | 99 | 9523 | 97% (166) | (2%) 9357 | 2% (166) | 98% (9357) |
| 99.5 | 5 | 0 | 0.3% (5) | 0 | 100% (5) | 99.5 | 2454 | 95% (163) | 0.5% (2291) | 7% (163) | 93% (2291) |
| 99.8 | 0 | - | - | - | - | 99.8 | 64 | 22% (38) | 0.005% (26) | 59% (38) | 41% (26) |
| - | - | - | - | - | - | 99.6 (Top 100) | 100 | 31% (53) | 0.01% (47) | 53% (53) | 47% (47) |

**Table S4:** Performance of FoundHaplo for the *SCN1B* c.363C>G disease haplotypes in the Epi25 and the UKBB cohorts. Since the total number of predictions in the UKBB at the $99^{th}$ percentile is too high for further screening, predictions were made by selecting the top 100 samples with the highest *FH* scores, assuming only 100 samples can be realistically screened in the UKBB cohort. The critical percentiles correspond to the percentiles of the 1000 Genomes control cohort. TP: true positive, FP: false positive, TD: True discovery and FD: false discovery.

| Epi25 | | | | | | UKBB | | | | | |
|---|---|---|---|---|---|---|---|---|---|---|---|
| **Critical percentile (%)** | **Samples predicted above the critical value** | **TP % (number)** | **FP % (number)** | **TD % (number)** | **FD % (number)** | **Critical percentile (%)** | **Samples predicted above the critical value** | **TP % (number)** | **FP % (number)** | **TD % (number)** | **FD % (number)** |
| 99 | 23 | 0 | 1.5% (23) | 0 | 100% (23) | 99 | 4459 | 97% (167) | 0.9% (4292) | 4% (167) | 96% (4292) |
| 99.5 | 6 | 0 | 0.4% (6) | 0 | 100% (6) | 99.5 | 2279 | 97% (167) | 0.45% (2112) | 7% (167) | 93% (2112) |
| 99.8 | 1 | 0 | 0.06% (1) | 0 | 100% (1) | 99.8 | 432 | 78% (134) | 0.06% (298) | 31% (134) | 69% (298) |
| - | - | - | - | - | - | 99.9 (Top 100) | 100 | 43% (74) | 0.005% (26) | 74% (74) | 26% (26) |

**Table S5:** Performance of FoundHaplo for the *WWOX* c.49G>A disease haplotypes in the Epi25 and the UKBB cohorts. Since the total number of predictions in the UKBB at the $99^{th}$ percentile is too high for further screening, predictions were made by selecting the top 100 samples with the highest *FH* scores, assuming only 100 samples can be realistically screened in the UKBB cohort. The critical percentiles correspond to the percentiles of the 1000 Genomes control cohort. TP: true positive, FP: false positive, TD: True discovery and FD: false discovery.

### Supplemental Material and Methods

#### FoundHaplo;
#### Software to predict individuals with disease-causing variants

FoundHaplo implements a first-order Hidden Markov Model (HMM) to infer the identity by descent (IBD) sharing between known disease haplotypes and individuals from cohorts to be screened with SNP genotyping array data (SNP chip data) for the existence of disease-causing variants (DCV) with known founder effects. Table S1 lists an illustrative set of inherited genetic disorders with reported founder effects.

FoundHaplo algorithm is implemented as an R package and is freely available from https://www.github.com/bahlolab/FoundHaplo.

For simplicity, we illustrate how the model is developed to test a single individual of interest using a single disease haplotype for a given DCV.

$H_0$ : The test individual does not share the DCV by IBD with the disease haplotype
$H_1$ : The test individual shares the DCV by IBD with the disease haplotype

This allows the detection of a DCV in either of the two test haplotypes. The algorithm does not extend to testing the number of copies of the DCV, which could be either one (present in only one haplotype) or two (present in both haplotypes).

If the FoundHaplo algorithm favours the null hypothesis ($H_0$), it doesn't definitively mean the test individual lacks the disease variant. Possible scenarios include: (i) the individual doesn't carry the DCV, (ii) the DCV is inherited from a different founder, i.e. has a different ancestral disease haplotype, or (iii) The pair has a very old common ancestor resulting in a small IBD segment (<=0.5 cM) indistinguishable from background linkage disequilibrium (LD) or constrained by the first-order Markov model's assumptions.

FoundHaplo computes the likelihood of IBD (denoted by 1) versus non-IBD (denoted by 0) using a log-likelihood ratio in the neighbourhood of the DCV for a given disease-test pair by using two Markov chains. Since the boundaries of the IBD sharing between disease and test individuals are not known in advance, we start the Markov chains at the DCV locus and traverse in opposing directions. L and R denote the ending points of the Markov chains to the left and right of the DCV locus, where the IBD segment around the DCV locus is inferred to have ended. The approach to determining the boundaries of the Markov chains is explained in subsequent sections.

Let $x$ be a disease haplotype known to harbour a genetic variant and $y$ a test individual of interest that we wish to test. Each test individual has two haplotypes, denoted by $h$ where $h \in H$, H={1,2}. Genetic markers in the FoundHaplo HMM are bi-allelic SNPs where the reference allele is denoted by 0 and the alternative allele by 1.

##### *Hidden state space*

The two Markov chains in the FoundHaplo algorithm are identical. Let $MC_L$ and $MC_R$ be the two Markov chains that traverse to the left and right, respectively, from the putative DCV locus. Negative integers denote the SNPs in the left chain, and positive integers denote the SNPs in the right chain. The DCV locus remains in between (t=0) the Markov chains and is shared by both as the starting state. We denote the sequence of hidden states in $MC_L$ by $Q_L = \{q_0^{x,y_h}, q_{-1}^{x,y_h}, q_{-2}^{x,y_h}, ..., q_L^{x,y_h}\}$ and in $MC_R$ by $Q_R = \{q_0^{x,y_h}, q_1^{x,y_h}, q_2^{x,y_h}, ..., q_R^{x,y_h}\}$.

##### *Initial probabilities*

Initial state probabilities vary based on the hypothesis and are denoted by $\Pi=\{\pi_1, \pi_2\}$. $\pi_1$ is defined as the probability of the Markov chain starting from a non-IBD state ($H_0$), and $\pi_2$ is defined as the probability of the Markov chain starting from an IBD state ($H_1$). Therefore, under $H_0$, $\pi_1$=1, $\pi_2$=0 and under $H_1$, $\pi_1$=0, $\pi_2$=1.

##### *Transition probabilities*

The Markov chain will transition from an IBD state to a non-IBD state if a recombination event occurs between the two markers. The number of crossovers between two genetic markers can be taken as being distributed according to a Poisson distribution with parameters $d_{i,j}$, where $d_{i,j}$ is the genetic map distance between loci $i$ and $j$ in Morgans [1,52]. The genetic map distances in FoundHaplo are taken from the HapMap project [53].

$$P(k \text{ crossovers in length } d_{i,j}) = \frac{\exp(-d_{i,j})d_{i,j}^k}{k!}$$
$$P(\text{no crossovers in length } d_{i,j}) = \exp(-d_{i,j})$$

Thus, the rate of transitioning from IBD to non-IBD is 1-exp($-d_{i,j}$), which denotes the probability of recombination between marker $i$ and $j$. The Markov chain will never be IBD again around the DCV once it reaches a non-IBD marker and is terminated at this point. Hence, the transition probability matrix, A, can be written as,

$$A = (a_{ij}) = \begin{pmatrix} 1 & 0 \\ 1 - \exp(-d_{i,j}) & \exp(-d_{i,j}) \end{pmatrix} \tag{1}$$

***Observation space***

At a given marker, there are four distinct pairwise allele combinations {(0,0),(0,1),(1,0)(1,1)} on a disease and test haplotype that makes up the observations. We denote the sequence of observations in $MC_L$ by $O_L = \{o_0^{x,y_h}, o_{-1}^{x,y_h}, o_{-2}^{x,y_h}, ..., o_L^{x,y_h}\}$ and in $MC_R$ by $O_R = \{o_0^{x,y_h}, o_1^{x,y_h}, o_2^{x,y_h}, ..., o_R^{x,y_h}\}$.

***Emission probabilities***

The emission probabilities $B$ of the Markov model are the probabilities of observing alleles given that they emit from a non-IBD or an IBD state.

***Emission probabilities*** $B$ ***with genotype and imputation errors***

Due to the potential for genotype and imputation errors in genetic data, the observed pairwise allele combinations may not match the true allele combinations.

There are four possible true pairwise allele combinations {(0,0),(0,1),(1,0)(1,1)} for a disease and a test haplotype at each marker. We denote the sequence of true alleles in $MC_L$ by $O'_L = \{o'^{x,y_h}_0, o'^{x,y_h}_{-1}, o'^{x,y_h}_{-2}, ..., o'^{x,y_h}_L\}$ and in $MC_R$ by $O'_R = \{o'^{x,y_h}_0, o'^{x,y_h}_1, o'^{x,y_h}_2, ..., o'^{x,y_h}_R\}$.

$$P(o'^{x}_t = 1) = P(o'^{y_h}_t = 1) = a_t$$
$$P(o'^{x}_t = 0) = P(o'^{y_h}_t = 0) = 1 - a_t$$

where, the probability of the alternate allele in disease or test haplotype is $a_t$, which is defined as the minor allele frequency (MAF) at marker $t$. MAF in the FoundHaplo algorithm is defined and annotated using population frequencies from the gnomAD public database [54,55].

The conditional probability of observing an allele on a haplotype given its true allele is,

$$P(o_t|o'_t) = \begin{cases} g, & \text{if } o_t \neq o'_t \\ 1 - g, & otherwise \end{cases} \tag{2}$$

The joint probability of the observed and true alleles is,

$$P(o_t, o\prime_t) = P(o_t | o\prime_t) P(o\prime_t) \tag{3}$$

Using equations 2 and 3, the genotype and imputation error probability $g$ is incorporated into the emission probability matrix $B$, as shown in Table S2.

The final transition ($A$) and emission probabilities ($B$) of the FoundHaplo HMM approach are further illustrated in Figure S1.

##### *Accommodating phasing errors in the model*

To accommodate phasing errors on the test individual, as the model traverses along the Markov chains, if the allele of the test haplotype of individual $y$ that the Markov chain is on does not match with the disease haplotype, the model switches to the other haplotype ($H \setminus$ h) looking for a match, as shown in algorithm S1.

**Algorithm S1:** Accommodating phasing errors in the algorithm.

**while** $t \geq L$ *if in* $MC_L$ *or* $t \leq R$ *if in* $MC_R$ **do**
  **if** $o_t^x \neq o_t^{y_h}$ *and* $o_t^x = o_t^{y_{H \setminus h}}$ **then**
    set $h = H \setminus$ h

##### *Calculating the FH score*

We define the likelihoods under the null and alternative hypotheses as $L_0$ and $L_1$ respectively, and define the FoundHaplo score, $FH$, for a single disease-test pair as the log of the log-likelihood ratio ($LLR$),

$$FH^{x,y} = \ln(LLR^{x,y}) \tag{4}$$

$$LLR^{x,y} = -2(\ln L_0^{x,y} - \ln L_1^{x,y}) \tag{5}$$

$L_0$ and $L_1$ are defined below with equations 6 and 7.

##### *Likelihood under the null hypothesis*

Under $H_0$, the Markov chain is non-IBD; hence we only consider the emission probabilities of $P(o_t^{x,y_h}, o\prime_t^{x,y_h} | q_t^{x,y_h} = 0)$ from $B$ shown in Table S2. We denote this subset of emission probabilities as $B_{H_0}$. The likelihood distribution under the null hypothesis is given by $L_0^{x,y}$ where,

$$
\begin{aligned}
L_0^{x,y} &= P(O, O\prime, Q|A, B_{H_0}, \Pi_{H_0}) \\
&= \prod_{t=-1}^{L} P(o_t^x|o\prime_t^x)P(o\prime_t^x)P(o_t^{y_h}|o\prime_t^{y_h})P(o\prime_t^{y_h}) \times \prod_{t=1}^{R} P(o_t^x|o\prime_t^x)P(o\prime_t^x)P(o_t^{y_h}|o\prime_t^{y_h})P(o\prime_t^{y_h}) \qquad (6)
\end{aligned}
$$

where, $h$ may swap as per algorithm S1.

*Likelihood under the alternative hypothesis*

Under $H_1$, we assume that the test individual shares the DCV by IBD, i.e. the test individual carries at least one copy of the disease allele. Hence, the initial state in the Markov chain is IBD. However, the Markov chain can transition to a non-IBD state at any marker. Therefore we have to consider both types of emission probabilities $P(o_t^{x,y_h}, o\prime_t^{x,y_h}|q_t^{x,y_h} = 0)$ and $P(o_t^{x,y_h}, o\prime_t^{x,y_h}|q_t^{x,y_h} = 1)$ from $B$, as shown in Table S2. We denote the set of emission probabilities that are used in $H_1$ as $B_{H_1}$, which is the total emission probability space of $B$. The likelihood distribution under the alternative hypothesis is given by $L_1^{x,y}$ where,

$$
\begin{aligned}
L_1^{x,y} &= P(O, O\prime, Q|A, B_{H_1}, \Pi_{H_1}) \\
&= \prod_{t=-1}^{L} [P(o\prime_t^x = o\prime_t^{y_h})P(o\prime_t^x)P(o_t^x|o\prime_t^x)P(o_t^{y_h}|o\prime_t^{y_h})\exp(-d_{t+1,t}) \\
&+ P(o_t^x|o\prime_t^x)P(o\prime_t^x)P(o_t^{y_h}|o\prime_t^{y_h})P(o\prime_t^{y_h})(1-\exp(-d_{t+1,t}))] \times \\
&\prod_{t=1}^{R} [P(o\prime_t^x = o\prime_t^{y_h})P(o\prime_t^x)P(o_t^x|o\prime_t^x)P(o_t^{y_h}|o\prime_t^{y_h})\exp(-d_{t-1,t}) \\
&+ P(o_t^x|o\prime_t^x)P(o\prime_t^x)P(o_t^{y_h}|o\prime_t^{y_h})P(o\prime_t^{y_h})(1-\exp(-d_{t-1,t}))] \qquad (7)
\end{aligned}
$$

where,

$$
P(o\prime_t^x = o\prime_t^{y_h}) = \begin{cases} 1, & \text{if } o\prime_t^x = o\prime_t^{y_h} \\ 0, & \text{otherwise} \end{cases}
$$

where, $h$ may swap as per algorithm S1.

##### *Stopping criteria of the Markov chains*

We need to determine the values of L and R where the Markov chain terminates. Ideally, L and R would correspond to the exact markers at which IBD sharing terminates. However, due to the possible presence of genotype, imputation and phasing errors, these cannot be determined precisely.

The stopping criteria for FoundHaplo were derived using a comprehensive simulation study using 1000 Genomes Phase 3 haplotypes [56]. According to the simulation results, Markov chains are terminated based on a heuristic $\Delta LLR_t^{x,y_h}$. We define $\Delta LLR_t^{x,y_h}$ as,

$$\Delta LLR_t^{x,y_h} = \begin{cases} LLR_t^{x,y_h} - LLR_{t+1}^{x,y_h}, & t < 0 \\ LLR_t^{x,y_h} - LLR_{t-1}^{x,y_h}, & t > 0 \end{cases}$$

$\Delta LLR_t^{x,y_h}$ is defined as the difference of $LLR$ values in the disease and test haplotypes between marker $t$ and the previous marker in the Markov chains. $LLR$ values can be calculated using equation 5.

We stop updating the Markov chain when the 100 points moving average of $\Delta LLR_t^{x,y_h}$ decreases to less than -0.4, which was chosen based on our simulation study. The endpoints of the Markov chains will be defined as markers L and R; these correspond to the endpoints of a potential IBD segment that carries the DCV. This is further discussed later in the simulation results in Figure S3.

##### *Incorporating multiple disease haplotypes*

$FH^y$ of a test sample $y$ when multiple known disease haplotypes are available for a given DCV is defined as,

$$LLR^{x,y} = -2(lnL_0^{x,y} - lnL_1^{x,y})$$
$$FH^{x,y} = ln(LLR^{x,y})$$
$$FH^y = \max_{x \in X} FH^{x,y}$$

where $X$ is the set of disease haplotypes available for a DCV.

We evaluated four different measures: summation($FH$), maximum($FH$), mean($FH$) and median($FH$) for combining $FH$ scores with multiple known disease haplotypes by comparing the Wilcoxon rank sum p-values for simulated cases versus controls. The maximum of individual $FH$ scores were selected as it differentiated simulated cases from controls the best. This is further discussed later in the simulation results in Figures S8-S9.

##### *Determining critical values to predict individuals that carry disease haplotypes*

The empirical null distribution of the *FH* scores from a suitable ancestral population of the 1000 Genomes haplotype data are used to determine the critical values in making predictions [56]. The default critical value is taken to be the $99^{th}$ percentile of the 1000 Genomes control cohort. Increasing the critical value percentile can reduce the empirical false positive rate. The predictions of the FoundHaplo algorithm in each analysis will be the test samples that gave *FH* score values higher than the *FH* score corresponding to the selected critical percentile in a control cohort.

#### Simulation study

##### *Simulating IBD segments*

The distances in Morgans from the DCV locus to the left or right breakpoints of the simulated ancestral segment in the descendant are distributed as independent exponential random variables with rate $n$, where $n$ is the number of generations since the most recent common ancestor MRCA [1]. Hence, the expected length of the left or right inherited ancestral segment in individuals who are $n^{th}$ generation descendants is $1/n$ Morgans, the mean of the exponential distribution. The total expected sharing lengths of the simulated IBD segments ($w$) between disease-case pairs in the simulation setup were set as 0.5, 1, 2 and 5 cM, sharing to either side corresponding to $w/2$. The number of generations ($n$) since the MRCA corresponding to the desired $w/2$ values can be estimated as $n = 100/(w/2)$. The left and right breakpoints in Figure S2 were, thus, stochastically generated using random exponential values with rate $n$. Since the simulated left and right breakpoints may differ, the simulated shared segments may not be symmetrical around the DCV locus.

##### *Simulation set up*

The 33 DCVs that we simulated to evaluate the performance of FoundHaplo are shown in Table S3. The left and right lengths of the shared haplotype between disease-case pairs were simulated as shown in Figure S2 for single and multiple founder effects. The entire process is repeated ten times to reflect ten different founder scenarios across the 33 DCVs, as shown in Algorithm S2.

**Algorithm S2:** Steps in the simulation set-up

**for** *seed* in 1 : 10 **do**
  **for** *DCV* in *DCV list with 33 DCVs* **do**
    **for** $w$ in $(0.5cM, 1cM, 2cM, 5cM)$ **do**
      Using $set.seed(seed)$, randomly generate,
      10 disease haplotypes
      50 (10 × 5) cases
      1 founder
      which gives 442 (503-10-50-1) controls in the simulated test cohort
      **if** Simulating single founder effect **then**
        **Perform step 1**
        Calculate the number of generations since the MRCA ($n$) →rexp(n) →Simulate IBD by splicing in a region of the core haplotype of the selected founder to the ten disease haplotypes
      **end if**
      **Perform step 2**
      Calculate the number of generations since the MRCA (n) based on $w$ →rexp(n) →Simulate IBD by splicing in a region of each disease haplotype to the respective five simulated cases
      **Add genotype/imputation and phasing errors**
      **Calculate** *FH* **score for**
      50 simulated cases using each of the 10 disease haplotypes
      442 controls in the simulated test cohort using each of the disease haplotype
      Save results for 4920 (50 × 10 + 442 × 10) comparisons
    **end for**
  **end for**
**end for**

##### *Deriving the stopping criteria of the Markov chains*

The Markov chains in the FoundHaplo algorithm must terminate after the IBD segment that carries the DCV has ended as the *FH* test statistic becomes progressively less powerful in discriminating between the null and alternative hypothesis. This stopping criteria of the Markov chain, after allowing for genotype/imputation and phasing errors, is derived based on the simulations.

Figure S3 represents an example of the behaviour of the *LLR* for a disease-case pair simulated to share 0.137 cM randomly generated length to the left, corresponding to 0.5

cM expected total sharing length (0.25 cM to the left), starting from the DCV locus to the left of the chromosome.

The *LLR* values keep increasing as long as the Markov chain is in an IBD segment that carries the DCV. After a while, the *LLR* starts to decrease when the alleles on both haplotypes of the test individual no longer share with the disease haplotype, and there are too many allele mismatches, signalling that the simulated IBD segment (0.137 cM, denoted by a solid red vertical line) has terminated. Based on the simulations, we end the Markov chains when the 100-marker moving average (calculated by taking 100 adjacent SNP markers with overlap) of the difference between adjacent *LLR* values decreases below -0.4 as shown in the green vertical line in Figure S3B. This threshold was found to be a good approximation for terminating the Markov chain based on simulation results across the 33 DCVs. Once the Markov chain is terminated, the algorithm traverses backwards in the chain, as shown in Figure S3C, to identify the point at which the Markov chain gave the highest *LLR* score. The corresponding genetic marker, shown in a dashed vertical red line in Figure S3C, is taken as the termination point in the Markov chain and as the length of the left arm of the shared IBD segment. The same approach is applied to the second Markov chain traversing to the right. The orange vertical line in Figure S3 C shows the point at which the Markov chain would have terminated if genotype/imputation and phasing errors were not accommodated. This point refers to the first allele mismatch between the disease-case pair. In this example, the first allele mismatch occurs after 0.0001 cM, whereas the true simulated sharing length is 0.137 cM. The Markov chain terminates very close to the starting point when errors are not accommodated. Figure S3 indicates that the FoundHaplo algorithm will likely overestimate the sharing length between two individuals when accommodating genotype/imputation and phasing errors due to the difficulty in determining the termination point of the two chains.

##### *Accuracy of the IBD segment length estimation in the FoundHaplo algorithm*

Haplotype sharing lengths between disease-case pairs are simulated in step 2 of the simulation setup. Ideally, the Markov chains in the algorithm would traverse and capture the exact respective simulated sharing length. However, the two Markov chains in the FoundHaplo algorithm could terminate before or after the simulated sharing length for simulated cases due to two reasons: (i) the algorithm failed to correctly predict the simulated cases because they were not distantly related to the disease haplotypes in use or too distantly related to the disease haplotypes, and (ii) the algorithm failed to correctly account for genotype/imputation and phasing errors. The Markov chain could terminate early on rare occasions where many genotype errors are clustered in a genomic region. Figure S4 shows the difference between the traversed length in cM in the Markov chain and the simulated sharing length drawn from a random exponential distribution using only the disease haplotypes and cases that were simulated to be distantly related for the (i) complete FoundHaplo algorithm that accommodates genotype/imputation and phasing errors and (ii) FoundHaplo algorithm without accommodating genotype/imputation and phasing errors. Using only the simulated distantly related disease-case pairs will isolate the portion of simulations that terminated the Markov chain before or after purely as a result of failing to account for genotype/imputation or phasing errors correctly.

The difference between the length traversed in the Markov chain and the simulated sharing length in Figure S4 for the complete FoundHaplo model has a mean of 0.4 cM and a standard error of 0.001. Only a small fraction of 1.73% of all the 132,000 (33 DCVs ×4 sharing lengths × 10 founder effect scenarios × 50 cases × 2 types of founder effects) simulations traversed less than the simulated length, and 98.27% of the simulations traversed more than the simulated length, with only 8% of simulations traversing more than 1 cM further from the simulated sharing length for the complete FoundHaplo model. This illustrates that the algorithm is likely to overestimate the length of haplotype sharing for a test individual. However, once the IBD segment has been traversed, any further genetic markers will dilute the evidence by decreasing the $FH$ statistic, which is also shown in Figure S3. Without accommodating genotype/imputation and phasing errors, less than 1% of total simulations traversed the simulated sharing length in the Markov chain.

##### *Using empirical p-values in the FoundHaplo algorithm*

Under the null hypothesis, the log-likelihood ratio test statistics are asymptotically chi-square distributed with degrees of freedom being the difference between the number of parameters in likelihoods under the null and alternative hypotheses [57].

Figure S5 shows the quantile-quantile plots of the $LLR$ values for controls. The X-axis represents the theoretical chi-squared distribution with degrees of freedom 1, and Y-axis represents the $LLR$ values for simulated controls which follow the null hypothesis.

Based on simulation results, a fraction of the controls gives very low $LLR$ scores and follows the theoretical chi-squared distribution with degrees of freedom 1 as shown in Figure S5. However, many controls deviate from the theoretical distribution ($-2(lnL_0^{x,y} - lnL_1^{x,y}) \not\equiv \chi_1$), giving moderate to high $LLR$ score values because they share the same disease haplotypes to different extents due to linkage disequilibrium or by chance. The sharing of controls for some example disease loci are shown in Figures S6-S7. We found that 5.38% of the simulated control individuals were detected as outliers (beyond the interquartile range) for the *LLR* values for single founder effects and 6.2% for multiple founder effects. Therefore, instead of using the theoretical chi-squared distribution, we chose to use the empirical distribution of *FH* scores to determine empirical p-values to make predictions.

The predictions of the FoundHaplo algorithm are made by selecting a critical value using the distribution of *FH* scores in the 1000 Genomes cohort as a control cohort. When using FoundHaplo, the control cohort should be of a similar ancestral population as the test cohort. This can be done by selecting the most relevant population of the five super populations (EUR, EAS, SAS, AMR and AFR) in the 1000 Genomes data. The control cohort in the simulations will be the EUR cohort of the 1000 Genomes data since simulations were performed using this cohort. The default critical value in FoundHaplo is taken as the $99^{th}$ percentile of the 1000 Genomes control cohort, allowing an empirical false positive rate of only 1%. Increasing the critical value percentile reduces the empirical false positive rate.

#### Detecting individuals with *SCN1B* c.363C>G and *WWOX* c.49G>A epilepsy variants

##### *Deriving the SCN1B c.363C>G and WWOX c.49G>A disease haplotypes*

The *SCN1B* c.363C>G (p.Cys121Trp) disease haplotypes were derived from a cohort of 100 individuals from nine Australian families recruited through the Epilepsy Research Centre, University of Melbourne. All the families were genotyped using the Illumina Global Screening Array-24. The *SCN1B* c.363C>G disease variant was confirmed in all nine families through Sanger sequencing or WES analysis [27]. Five *SCN1B* c.363C>G disease haplotypes were derived by pedigree phasing affected offspring of five families.

The *WWOX* c.49G>A (p.E17K) disease haplotypes were derived from a cohort of three duos recruited through the Epilepsy Research Centre, University of Melbourne. All the families were genotyped using the Illumina Global Screening Array-24. The *WWOX* c.49G>A disease variant was confirmed in all six individuals through WES. Three *WWOX* c.49G>A disease haplotypes were derived by pedigree phasing affected offspring of the three duos.

Both *SCN1B* c.363C>G and *WWOX* c.49G>A families were of European ancestry. When creating disease haplotypes, samples with a call rate of less than 98% and SNPs with an overall call rate of less than 98% were removed as initial quality control steps. The data were harmonised to 1000 Genomes data using Genotype Harmonizer [58]. Before imputation and phasing, the resulting data were converted to VCF format (Variant call format) and the imputation was performed using the Michigan Imputation Server (MIS) [59] with the European cohort of the 1000 Genomes phase 3 haplotypes (hg19 human genome build) [56] as the reference panel. Minor allele frequencies were annotated from the gnomAD population frequencies [54,55]. The data set was trimmed to contain a 20 cM region around the DCV with 10 cM on either side of the respective DCV loci. (chr19:35524558 and chr16:78133724 for the *SCN1B* c.363C>G and *WWOX* c.49G>A variants respectively). Only biallelic markers with a p-value for Hardy–Weinberg equilibrium chi-squared test $\geq 0.0000001$ and $R^2$ (imputation quality score) $\geq 0.3$ were retained. Since the two resulting data sets contain individuals of the same family with the *SCN1B* c.363C>G and *WWOX* c.49G>A variants, they were pedigree phased to extract five disease haplotypes for *SCN1B* c.363C>G variant and three disease haplotype for *WWOX* c.49G>A variant.

##### *Epi25 test cohort (n=1,573)*

The Epi25 test cohort consists of 1,573 individuals with different forms of epilepsy recruited in Australia and New Zealand and sequenced as part of the Epi25 Collaborative [60]. The Epi25 samples are both genotyped and whole exome sequenced. Genotyping was performed using the Illumina Infinium Global Screening Array with multi-disease content (GSA-MD v1.0) [60]. Chromosomes 19 and 16 of the Epi25 test cohort were prepared following the same process as in preparing the *SCN1B* c.363C>G and *WWOX* c.49G>A disease cohorts.

##### *UKBB test cohort (n=468,481)*

The UKBB cohort that was analysed using FoundHaplo consisted of 468,481 samples with both imputed SNP genotyping and WES data. The data was accessed through project ID 36610 (PI Bahlo). As the UKBB cohort was already imputed, samples only needed to be phased to run FoundHaplo, which was performed in-house using SHAPEIT4 (version 4.2.2) [61].

Imputed SNP genotyping data was converted from bgen format to VCF format using PLINK [62], and samples withdrawn from the UKBB project were removed. Chromosomes 19 and 16 were then phased using SHAPEIT4 (version 4.2.2) with the hg19 human genome build. No reference panel was provided. After phasing, only biallelic markers with a p-value for Hardy–Weinberg equilibrium chi-squared test $\geq 0.0000001$ and $R^2$ (imputation quality score) $\geq 0.3$ were retained. The data set was trimmed to contain only a 20 cM region around the *SCN1B* c.363C>G and *WWOX* c.49G>A variants with 10 cM on either side.

##### *Comparing FoundHaplo performance with DRIVE*

The DRIVE algorithm [63] was used to detect carriers of *SCN1B* c.363C>G and *WWOX* c.49G>A variants in the Epi25 cohort using the constructed disease haplotypes. We merged known *SCN1B* c.363C>G and *WWOX* c.49G>A DCV carriers into the Epi25 dataset in two separate analyses to determine whether other individuals with these DCVs in the Epi25 cohort are clustered together with the already known DCV carriers. We did not run DRIVE on the UKBB cohort as the required run time given the size of this cohort was not feasible.

Only biallelic markers with MAF >0.01, markers with a p-value for Hardy–Weinberg equilibrium chi-squared test $\geq 0.0000001$ and $R^2$ (imputation quality score) $\geq 0.3$ were used in the analysis. The target region for each DCV was set as 0.5 cM around the DCV locus, with 0.25 cM on either side. We used the default minimum IBD length of 3 cM (min-cm argument in DRIVE) as specified by the DRIVE tool.

The DRIVE tool filters out segments that don't contain the entire target region of 0.5 cM around the DCV loci and all pairwise IBD segments that are shorter than the provided minimum length threshold of 3 cM. Pairwise IBD segments were generated using hap-ibd by Zhou et al. [64]. The carriers were detected based on clustering with at least two of the known disease haplotypes.

##### *Haplotype Analysis*

Imputed and phased *SCN1B* c.363C>G and *WWOX* c.49G>A disease haplotypes were filtered to retain markers with imputation accuracy $\geq$0.9. These haplotypes around the two disease variants were compared to evaluate their sharing with each other. The haplotypes carrying the *SCN1B* c.363C>G and *WWOX* c.49G>A variants in the Epi25 and the UKBB cohorts were then compared with each of the disease haplotypes. Only the markers with imputation accuracy $\geq$0.9 were retained in the analysis (Figures S10-S11).

##### *Ethics Statement*

This study was approved by the Austin Health Human Research Ethics Committee. Informed consent was obtained and archived from all participants or their legal guardian. Research was approved by the Human Research Ethics Committee at The Walter and Eliza Hall Institute of Medical Research (G20/01, 17/09LR).

##### *Supplemental Acknowledgements*

We thank the Epi25 principal investigators, local staff from individual cohorts, and the individuals with epilepsy who participated in Epi25 for making possible this global collaboration and resource to advance epilepsy genetics research. This research was conducted with data from UK Biobank (www.ukbiobank.ac.uk), a major biomedical database, under data use agreement 36610 (PI Bahlo).

**Supplemental References**


1. McPeek, M. S. and Strahs, A. (1999). Assessment of linkage disequilibrium by the decay of haplotype sharing, with application to fine-scale genetic mapping. Am. J. Hum. Genet. *65*, 858–875.

2. Rafehi, H., Szmulewicz, D. J., Bennett, M. F., Sobreira, N. L. M., Pope, K., Smith, K. R., Gillies, G., Diakumis, P., Dolzhenko, E., Eberle, M. A., et al. (2019). Bioinformatics-Based identification of expanded repeats: A non-reference intronic pentamer expansion in RFC1 causes CANVAS. Am. J. Hum. Genet. *105*, 151–165.

3. Cortese, A., Simone, R., Sullivan, R., Vandrovcova, J., Tariq, H., Yan, Y. W., Humphrey, J., Jaunmuktane, Z., Sivakumar, P., Polke, J., et al. (2019). Biallelic expansion of an intronic repeat in RFC1 is a common cause of late-onset ataxia. Nat. Genet. *51*, 649–658.

4. Mitchison, H. M., O'Rawe, A. M., Taschner, P. E., Sandkuijl, L. A., Santavuori, P., de Vos, N., Breuning, M. H., Mole, S. E., Gardiner, R. M., and Järvelä, I. E. (1995). Batten disease gene, CLN3: linkage disequilibrium mapping in the finnish population, and analysis of european haplotypes. Am. J. Hum. Genet. *56*, 654–662.

5. Morral, N., Bertranpetit, J., Estivill, X., Nunes, V., Casals, T., Gimenez, J., Reis, A., Varon-Mateeva, R., Macek Jr, M., Kalaydjieva, L., et al. (1994). The origin of the major cystic fibrosis mutation ($\delta$f508) in european populations. Nature genetics *7*, 169–175.

6. Martins, S., Matamá, T., Guimarães, L., Vale, J., Guimarães, J., Ramos, L., Coutinho, P., Sequeiros, J., and Silveira, I. (2003). Portuguese families with dentatorubropallidoluysian atrophy (DRPLA) share a common haplotype of asian origin. Eur. J. Hum. Genet. *11*, 808–811.

7. Grimaldi, S., Cupidi, C., Smirne, N., Bernardi, L., Giacalone, F., Piccione, G., Basiricò, S., Mangano, G. D., Nardello, R., Orsi, L., et al. (2019). The largest caucasian kindred with dentatorubral-pallidoluysian atrophy: A founder mutation in italy. Mov. Disord. *34*, 1919–1924.

8. Weisz-Hubshman, M., Meirson, H., Michaelson-Cohen, R., Beeri, R., Tzur, S., Bormans, C., Modai, S., Shomron, N., Shilon, Y., Banne, E., et al. (2019). Novel WWOX deleterious variants cause early infantile epileptic encephalopathy, severe developmental delay and dysmorphism among yemenite jews. Eur. J. Paediatr. Neurol. *23*, 418–426.

9. Ishiura, H., Ishiura, H., Doi, K., Doi, K., Mitsui, J., Mitsui, J., Yoshimura, J., Yoshimura, J., Matsukawa, M. K., Matsukawa, M. K., et al. (2018). Expansions of intronic TTTCA and TTTTA repeats in benign adult familial myoclonic epilepsy. Nat. Genet. *50*, 581–590.

10. Cen, Z., Jiang, Z., Chen, Y., Zheng, X., Xie, F., Yang, X., Lu, X., Ouyang, Z., Wu, H., Chen, S., et al. (2018). Intronic pentanucleotide TTTCA repeat insertion in the SAMD12 gene causes familial cortical myoclonic tremor with epilepsy type 1. Brain *141*, 2280–2288.

11. Bennett, M. F., Oliver, K. L., Regan, B. M., Bellows, S. T., Schneider, A. L., Rafehi, H., Sikta, N., Crompton, D. E., Coleman, M., Hildebrand, M. S., et al. (2020). Familial adult myoclonic epilepsy type 1 SAMD12 TTTCA repeat expansion arose 17,000 years ago and is present in sri lankan and indian families. Eur. J. Hum. Genet. *28*, 973–978.

12. Yeetong, P., Chunharas, C., Pongpanich, M., Bennett, M. F., Srichomthong, C., Pasutharnchat, N., Suphapeetiporn, K., Bahlo, M., and Shotelersuk, V. (2020). Founder effect of the TTTCA repeat insertions in SAMD12 causing BAFME1. Eur. J. Hum. Genet.

13. Madia, F., Striano, P., Di Bonaventura, C., de Falco, A., de Falco, F. A., Manfredi, M., Casari, G., Striano, S., Minetti, C., and Zara, F. (2008). Benign adult familial myoclonic epilepsy (BAFME): evidence of an extended founder haplotype on chromosome 2p11.1-q12.2 in five italian families. Neurogenetics *9*, 139–142.

14. Henden, L., Freytag, S., Afawi, Z., Baldassari, S., Berkovic, S. F., Bisulli, F., Canafoglia, L., Casari, G., Crompton, D. E., Depienne, C., et al. (2016). Identity by descent fine mapping of familial adult myoclonus epilepsy (FAME) to 2p11.2-2q11.2. Hum. Genet. *135*, 1117–1125.

15. Florian, R. T., Kraft, F., Leitão, E., Kaya, S., Klebe, S., Magnin, E., van Rootselaar, A.-F., Buratti, J., Kühnel, T., Schröder, C., et al. (2019). Unstable TTTTA/TTTCA expansions in MARCH6 are associated with familial adult myoclonic epilepsy type 3. Nat. Commun. *10*, 4919.

16. Kengne Kamga, K., Nguefack, S., Minka, K., Wonkam Tingang, E., Esterhuizen, A., Nchangwi Munung, S., De Vries, J., and Wonkam, A. (2020). Cascade testing for fragile X syndrome in a rural setting in cameroon (Sub-Saharan africa). Genes *11*.

17. Oudet, C., von Koskull, H., Nordström, A. M., Peippo, M., and Mandel, J. L. (1993). Striking founder effect for the fragile X syndrome in finland. Eur. J. Hum. Genet. *1*, 181–189.

18. Malmgren, H., Gustavson, K. H., Oudet, C., Holmgren, G., Pettersson, U., and Dahl, N. (1994). Strong founder effect for the fragile X syndrome in sweden. Eur. J. Hum. Genet. *2*, 103–109.

19. Cossée, M., Schmitt, M., Campuzano, V., Reutenauer, L., Moutou, C., Mandel, J. L., and Koenig, M. (1997). Evolution of the friedreich's ataxia trinucleotide repeat expansion: founder effect and premutations. Proc. Natl. Acad. Sci. U. S. A. *94*, 7452–7457.

20. Monrós, E., Cañizares, J., Moltó, M. D., Rodius, F., Montermini, L., Cossée, M., Martínez, F., Prieto, F., de Frutos, R., Koenig, M., et al. (1996). Evidence for a common origin of most friedreich ataxia chromosomes in the spanish population. Eur. J. Hum. Genet. *4*, 191–198.

21. Sirugo, G., Keats, B., Fujita, R., Duclos, F., Purohit, K., Koenig, M., and Mandel, J. L. (1992). Friedreich ataxia in louisiana acadians: demonstration of a founder effect by analysis of microsatellite-generated extended haplotypes. Am. J. Hum. Genet. *50*, 559–566.

22. Zühlke, C., Laccone, F., Cossée, M., Kohlschütter, A., Koenig, M., and Schwinger, E. (1998). Mutation of the start codon in the FRDA1 gene: linkage analysis of three pedigrees with the ATG to ATT transversion points to a unique common ancestor. Hum. Genet. *103*, 102–105.

23. Smith, B. N., Newhouse, S., Shatunov, A., Vance, C., Topp, S., Johnson, L., Miller, J., Lee, Y., Troakes, C., Scott, K. M., et al. (2013). The C9ORF72 expansion mutation is a common cause of ALS+/FTD in europe and has a single founder. Eur. J. Hum. Genet. *21*, 102–108.

24. Mok, K., Traynor, B. J., Schymick, J., Tienari, P. J., Laaksovirta, H., Peuralinna, T., Myllykangas, L., Chiò, A., Shatunov, A., Boeve, B. F., et al. (2012). Chromosome 9 ALS and FTD locus is probably derived from a single founder. Neurobiol. Aging *33*, 209.e3–8.

25. Laaksovirta, H., Peuralinna, T., Schymick, J. C., Scholz, S. W., Lai, S.-L., Myllykangas, L., Sulkava, R., Jansson, L., Hernandez, D. G., Gibbs, J. R., et al. (2010). Chromosome

9p21 in amyotrophic lateral sclerosis in finland: a genome-wide association study. Lancet Neurol. *9*, 978–985.

26. Ishiura, H., Takahashi, Y., Mitsui, J., Yoshida, S., Kihira, T., Kokubo, Y., Kuzuhara, S., Ranum, L. P. W., Tamaoki, T., Ichikawa, Y., et al. (2012). C9ORF72 repeat expansion in amyotrophic lateral sclerosis in the kii peninsula of japan. Arch. Neurol. *69*, 1154–1158.

27. Grinton, B. E., Robertson, E., Fearnley, L. G., Scheffer, I. E., Marson, A. G., O'Brien, T. J., Pickrell, W. O., Rees, M. I., Sisodiya, S. M., Balding, D. J., et al. (2022). A founder event causing a dominant childhood epilepsy survives 800 years through weak selective pressure. Am. J. Hum. Genet. *109*, 2080–2087.

28. Paradisi, I., Hernández, A., and Arias, S. (2008). Huntington disease mutation in venezuela: age of onset, haplotype analyses and geographic aggregation. J. Hum. Genet. *53*, 127–135.

29. Nakashima, K., Watanabe, Y., Kusumi, M., Nanba, E., Maeoka, Y., Igo, M., Irie, H., Ishino, H., Fujimoto, A., and Kobayashi, S. (1995). [prevalence and founder effect of huntington's disease in the san-in area of japan]. Rinsho Shinkeigaku *35*, 1532–1534.

30. García-Planells, J., Burguera, J. A., Solís, P., Millán, J. M., Ginestar, D., Palau, F., and Espinós, C. (2005). Ancient origin of the cag expansion causing huntington disease in a spanish population. Human mutation *25*, 453–459.

31. Demetriou, C., Heraclides, A., Salafori, C., Tanteles, G., Christodoulou, K., Christou, Y., and Zamba-Papanicolaou, E. (2018). Epidemiology of huntington disease in cyprus: A 20-year retrospective study. Clinical Genetics *93*, 656–664.

32. Yotova, V., Labuda, D., Zietkiewicz, E., Gehl, D., Lovell, A., Lefebvre, J.-F., Bourgeois, S., Lemieux-Blanchard, E., Labuda, M., Vézina, H., et al. (2005). Anatomy of a founder effect: myotonic dystrophy in northeastern quebec. Hum. Genet. *117*, 177–187.

33. Goldman, A., Krause, A., Ramsay, M., and Jenkins, T. (1996). Founder effect and prevalence of myotonic dystrophy in south africans: molecular studies. Am. J. Hum. Genet. *59*, 445–452.

34. Moulard, B., Darcel, F., Mignard, D., Jeanpierre, M., Genton, P., Cartault, F., Yaouanq, J., Roubertie, A., Biraben, A., Buresi, C., et al. (2003). FOunder effect in patients with Unverricht-Lundborg disease on reunion island. Epilepsia *44*, 1357–1360.

35. Boissé Lomax, L., Bayly, M. A., Hjalgrim, H., Møller, R. S., Vlaar, A. M., Aaberg, K. M., Marquardt, I., Gandolfo, L. C., Willemsen, M., Kamsteeg, E.-J., et al. (2013). 'north sea' progressive myoclonus epilepsy: phenotype of subjects with GOSR2 mutation. Brain *136*, 1146–1154.

36. Tanaka, F., Doyu, M., Ito, Y., Matsumoto, M., Mitsuma, T., Abe, K., Aoki, M., Itoyama, Y., Fischbeck, K. H., and Sobue, G. (1996). Founder effect in spinal and bulbar muscular atrophy (SBMA). Hum. Mol. Genet. *5*, 1253–1257.

37. Lund, A., Udd, B., Juvonen, V., Andersen, P. M., Cederquist, K., Davis, M., Gellera, C., Kölmel, C., Ronnevi, L. O., Sperfeld, A. D., et al. (2001). Multiple founder effects in spinal and bulbar muscular atrophy (SBMA, kennedy disease) around the world. Eur. J. Hum. Genet. *9*, 431–436.

38. Lund, A., Udd, B., Juvonen, V., Andersen, P. M., Cederquist, K., Ronnevi, L.-O., Sistonen, P., Sörensen, S. A., Tranebjærg, L., Wallgren-Pettersson, C., et al. (2000). Founder effect in spinal and bulbar muscular atrophy (SBMA) in scandinavia. Eur. J. Hum. Genet. *8*, 631–636.

39. Pérez, L. V., Cruz, G. S., Falcón, N. S., Mederos, L. E. A., Batallan, K. E., Labrada, R. R., Herrera, M. P., Mesa, J. M. L., Díaz, J. C. R., Rodríguez, R. A., et al. (2009). Molecular epidemiology of spinocerebellar ataxias in cuba: insights into sca2 founder effect in holguin. Neuroscience letters *454*, 157–160.

40. Mizushima, K., Watanabe, M., Kondo, I., Okamoto, K., Shizuka, M., Abe, K., Aoki, M., and Shoji, M. (1999). Analysis of spinocerebellar ataxia type 2 gene and haplotype analysis:(ccg) 1-2 polymorphism and contribution to founder effect. Journal of medical genetics *36*, 112–114.

41. Stevanin, G., Cancel, G., Didierjean, O., Dürr, A., Abbas, N., Cassa, E., Feingold, J., Agid, Y., and Brice, A. (1995). Linkage disequilibrium at the Machado-Joseph disease/spinal cerebellar ataxia 3 locus: evidence for a common founder effect in french and Portuguese-Brazilian families as well as a second ancestral Portuguese-Azorean mutation. Am. J. Hum. Genet. *57*, 1247–1250.

42. Li, T., Martins, S., Peng, Y., Wang, P., Hou, X., Chen, Z., Wang, C., Tang, Z., Qiu, R., Chen, C., et al. (2018). Is the high frequency of Machado-Joseph disease in china due to new mutational origins? Front. Genet. *9*, 740.

43. Mori, M., Adachi, Y., Kusumi, M., and Nakashima, K. (2001). Spinocerebellar ataxia type 6: founder effect in western japan. J. Neurol. Sci. *185*, 43–47.

44. Dichgans, M., Schöls, L., Herzog, J., Stevanin, G., Weirich-Schwaiger, H., Rouleau, G., Bürk, K., Klockgether, T., Zühlke, C., Laccone, F., et al. (1999). Spinocerebellar ataxia

type 6: evidence for a strong founder effect among german families. Neurology *52*, 849–851.

45. Almeida, T., Alonso, I., Martins, S., Ramos, E. M., Azevedo, L., Ohno, K., Amorim, A., Saraiva-Pereira, M. L., Jardim, L. B., Matsuura, T., et al. (2009). Ancestral origin of the ATTCT repeat expansion in spinocerebellar ataxia type 10 (SCA10). PLoS One *4*, e4553.

46. Bushara, K., Bower, M., Liu, J., McFarland, K. N., Landrian, I., Hutter, D., Teive, H. A. G., Rasmussen, A., Mulligan, C. J., and Ashizawa, T. (2013). Expansion of the spinocerebellar ataxia type 10 (SCA10) repeat in a patient with sioux native american ancestry. PLoS One *8*, e81342.

47. Bahl, S., Virdi, K., Mittal, U., Sachdeva, M. P., Kalla, A. K., Holmes, S. E., O'Hearn, E., Margolis, R. L., Jain, S., Srivastava, A. K., et al. (2005). Evidence of a common founder for SCA12 in the indian population. Ann. Hum. Genet. *69*, 528–534.

48. Zühlke, C., Dalski, A., Schwinger, E., and Finckh, U. (2005). Spinocerebellar ataxia type 17: report of a family with reduced penetrance of an unstable gln49 TBP allele, haplotype analysis supporting a founder effect for unstable alleles and comparative analysis of SCA17 genotypes. BMC Med. Genet. *6*, 27.

49. Amino, T., Ishikawa, K., Toru, S., Ishiguro, T., Sato, N., Tsunemi, T., Murata, M., Kobayashi, K., Inazawa, J., Toda, T., et al. (2007). Redefining the disease locus of 16q22.1-linked autosomal dominant cerebellar ataxia. J. Hum. Genet. *52*, 643–649.

50. Hirano, R., Takashima, H., Okubo, R., Tajima, K., Okamoto, Y., Ishida, S., Tsuruta, K., Arisato, T., Arata, H., Nakagawa, M., et al. (2004). Fine mapping of 16q-linked autosomal dominant cerebellar ataxia type III in japanese families. Neurogenetics *5*, 215–221.

51. Takashima, M., Ishikawa, K., Nagaoka, U., Shoji, S., and Mizusawa, H. (2001). A linkage disequilibrium at the candidate gene locus for 16q-linked autosomal dominant cerebellar ataxia type III in japan. J. Hum. Genet. *46*, 167–171.

52. S., H. J. B. (1919). The combination of linkage values and the calculation of distance between the loci fo linked factors. Hournal of Gnetics *8*, 299–309.

53. International HapMap Consortium. (2003). The international HapMap project. Nature *426*, 789–796.

54. Karczewski, K. J., Francioli, L. C., Tiao, G., Cummings, B. B., Alföldi, J., Wang, Q., Collins, R. L., Laricchia, K. M., Ganna, A., Birnbaum, D. P., et al. (2020).

The mutational constraint spectrum quantified from variation in 141,456 humans. Nature *581*, 434–443.

55. Chen, S., Francioli, L. C., Goodrich, J. K., Collins, R. L., Kanai, M., Wang, Q., Alföldi, J., Watts, N. A., Vittal, C., Gauthier, L. D., et al. (2023). A genomic mutational constraint map using variation in 76,156 human genomes. Nature pp. 1–11.

56. 1000 Genomes Project Consortium, Auton, A., Brooks, L. D., Durbin, R. M., Garrison, E. P., Kang, H. M., Korbel, J. O., Marchini, J. L., McCarthy, S., McVean, G. A., et al. (2015). A global reference for human genetic variation. Nature *526*, 68–74.

57. Wilks, S. S. (1938). The Large-Sample distribution of the likelihood ratio for testing composite hypotheses. Ann. Math. Stat. *9*, 60–62.

58. Deelen, P., Bonder, M. J., van der Velde, K. J., Westra, H.-J., Winder, E., Hendriksen, D., Franke, L., and Swertz, M. A. (2014). Genotype harmonizer: automatic strand alignment and format conversion for genotype data integration. BMC Res. Notes *7*, 901.

59. Das, S., Forer, L., Schönherr, S., Sidore, C., Locke, A. E., Kwong, A., Vrieze, S. I., Chew, E. Y., Levy, S., McGue, M., et al. (2016). Next-generation genotype imputation service and methods. Nat. Genet. *48*, 1284–1287.

60. Epi25 Collaborative. Electronic address: and Epi25 Collaborative. (2019). Ultra-Rare genetic variation in the epilepsies: A Whole-Exome sequencing study of 17,606 individuals. Am. J. Hum. Genet. *105*, 267–282.

61. Delaneau, O., Coulonges, C., and Zagury, J.-F. (2008). Shape-IT: new rapid and accurate algorithm for haplotype inference. BMC Bioinformatics *9*, 540.

62. Purcell, S., Neale, B., Todd-Brown, K., Thomas, L., Ferreira, M. A. R., Bender, D., Maller, J., Sklar, P., de Bakker, P. I. W., Daly, M. J., et al. (2007). PLINK: a tool set for whole-genome association and population-based linkage analyses. Am. J. Hum. Genet. *81*, 559–575.

63. Lancaster, M. C., Chen, H.-H., Shoemaker, M. B., Fleming, M. R., Baker, J. T., Polikowsky, H. G., Samuels, D. C., Huff, C. D., Roden, D. M., and Below, J. E. (2023). Detection of distant familial relatedness in biobanks for identification of undiagnosed carriers of a mendelian disease variant: application to long qt syndrome. medRxiv pp. 2023–04.

64. Zhou, Y., Browning, S. R., and Browning, B. L. (2020). A fast and simple method for detecting Identity-by-Descent segments in Large-Scale data. Am. J. Hum. Genet. *106*, 426–437.
